# Identification of Blood Glucose Patterns through Continuous Glucose Monitoring Sensors and Decision Trees

**DOI:** 10.1101/2020.09.09.20190736

**Authors:** F. José Lozano, J. Ignacio Hidalgo, Marta Botella, Sergio Contador, Juan Lanchares, J. Manuel Velasco, Oscar Garnica

## Abstract

The demand for Continuous Glucose Monitoring systems is increasing among type 1 diabetic patients. Some companies are trying to improve the monitoring and the usability of these systems. One example is Abbott FreeStyle Libre, which provides a new concept of glucose monitoring called Flash Glucose Monitoring which is more affordable and does not need calibration. The increasing demand for these devices means an opportunity for data and computer scientists, who can contribute to the development of decision-making support systems based on the data collected from the devices. Type 1 diabetic patients that use FreeStyle Libre may enter the number of insulin and carbohydrates units that they are going to take before a meal. Using both the entered data and the blood glucose values collected by the device automatically, the application presented in this paper generates a report of the patient’s glucose patterns. In addition, it provides a web application that allows the user to upload the data obtained from the device and download the report on his computer or smartphone. The application uses decision trees to detect the patterns and entails a starting point in the creation of ensemble models with more predictive power, also based on decision trees. Furthermore, the methodology makes a segmentation of the dataset in blocks, determined by the different meals done throughout the day, adding more information to the set of variables used to train the model. As a result, the application can discover repetitive patterns in the daily life of the patient, which can help to take early preventive measures for risk situations in a period close to the next meal.

## 1 Introduction

Diabetes mellitus is a chronic disease that is characterised by the presence of hyperglycemia as a consequence of defects in insulin secretion, insulin action or both [1]. Diabetes mellitus can be mainly classified into four different types:

- Type 1 diabetes mellitus, which causes the destruction of a beta cell that is present in the pancreas, and an absolute deficiency in insulin production.
- Type 2 diabetes mellitus, which causes resistance to the insulin action and certain deficiency in the insulin secretion.
- Other types of diabetes, related to genetic defects that affect to the beta cell such as pancreatic lesions, drugs and other factors.
- Gestational diabetes, which appears during the gestation period.

It is estimated five percent of the Spanish population suffers from type 1 diabetes mellitus [2]. Although this kind of diabetes is not related to the lifestyle, its incidence is increasing among the population. Diabetics must take an insulin dose before or shortly after each meal to keep their blood glucose level in harmless range. Many tools allow diabetic patients to monitor their blood glucose values in real time and have a better control of their disease. The increment in the commercialization of blood glucose monitoring systems is an opportunity for data and computer scientists to create innovative tools that help diabetics to anticipate to upcoming risk situations and improve their lifestyle. It is important to provide them with a service that detect repetitive risky situations, but it is even more important to give them meaningful answers according to these patterns. This last guidance must be by thoroughly designed by means of a joint effort among computer scientist and physicians, who are responsible for giving a correct characterization to the identified patterns.

This work aims to provide a decision-making support system to type 1 diabetic patients. The application uses the data obtained from a Flash Glucose Monitoring (FGM) device. The main difference with traditional Continuous Glucose Monitoring (CGM) devices is that the patient does not see his blood sugar levels until he scans the sensor with the reader. Another difference is that the device does not need to be calibrated while some other CGM devices need to be calibrated several times per day [3].

The patient has access to a report of patterns of his blood glucose values by uploading the data of the reader to a web application. This idea has been inspired by other studies and applications focused on the development of a solution for diabetic people using data from a monitoring system. One example of an application developed for diabetics is glUCModel, a monitoring and modelling system for chronic diseases applied to diabetes [4]. This application allows doctors to consult the information of their patients and have better control over their illness. Moreover, glUCModel offers a recommendation system that provides automatic recommendations to their patients, increasing their awareness of their condition. This project has a similar goal, but using a different approach through decision trees to recognise the patterns and segmenting each day in blocks determined by the meals.

Another study that has provided some ideas to this project has been described in [5]. This thesis presents a support system to diabetics using data from a CGM device and machine learning techniques, which try to predict the blood glucose values of the patients. This study contains interesting contributions about data preprocessing, feature engineering, classification of glycemic variability and evaluation techniques.

Currently, many different models try to predict the blood glucose values of diabetic patients and warn them when these values are not in a safe range. However, these models often act like black boxes that send alerts, but do not provide an explanation regarding their provenance. Furthermore, the patient sometimes has a greater need of knowing how to react to frequent dangerous situations that compromise their health. Once the patient becomes aware of these patterns, the patient would be able to modify their daily habits to avoid unwanted scenarios.

Detecting patterns in blood glucose levels is a challenge that we wanted to face with this work. Giving useful and accessible descriptions to both patients and physicians is even more challenging. It is necessary to innovate to address these challenges and provide solutions potentially helpful. However, innovation and willingness are not sufficient if they are not coupled with the creation of realistic scenarios using actual data. Working with data collected from diabetic patients is something that made this project very attractive to us because accessing to real information is often limited or restricted.

Data-driven solutions are a trend in the last years due to the existence of tens of data mining and big data tools to discover hidden patterns in large datasets. This project was an opportunity to turn our ideas into action and use our knowledge in data science to improve the quality life of diabetics. Further, we wanted to contribute to the research of new methods of prevention of hyperglycemia. We believe that computer scientists have the social responsibility of providing solutions to society problems using the available technology. Developing this application is only one small contribution to a collective effort of helping diabetics to live a life as normal as possible.

In this paper we developed a Python package that generates reports using data files obtained from a FreeStyle Libre sensor, a blood glucose monitoring device. We also developed a web application that integrated the package and offered its services to the user through a web browser. The main package is not intended to be a predictive tool for the patient but rather a descriptive instrument to support decision-making. The application is not connected in real time to another device or smartphone, but it uses the data files obtained by the patient from the reader. Despite these limitations, another objective of the application is still being useful for the patient.

The rest of the paper is structured as follows. In section 2, we describe the materials and methods that allowed us to work on this project. Firstly, the device in charge of monitoring the blood glucose values and secondly, the decision trees used to create the model, explaining their features and the algorithm they use. Section 3 explains the design of both the application core and the web application that wraps the core. Section 4 describes the whole methodology from the beginning, when the application reads the data files and process them, to the creation of the model and the generation of the report. Section 5 shows the results of applying the methodology in a series of data files, commenting the relevant ones and discussing which were expected beforehand. Finally, section 6 contains a conclusion and suggests ideas for a possible improvement of the methodology.

## 2 Materials and methods

### 2.1 FreeStyle Blood Glucose Monitoring System

The data files that are processed in this web are obtained from FreeStyle Libre devices, manufactured by the company Abbott Diabetes Care, which sells several appliances for glucose control by diabetic patients. CGM became available in the year 2000, having a measurement error of more than *±* 20 %. New devices have reduced this error to 10 %, but there is a continuous effort for reducing this margin by the producers [6].

FreeStyle Libre is an interstitial CGM system, which is an alternative for the capillary and venous blood glucose measurement [7]. One advantage of this kind of devices is that provides more reference points of measurement than conventional blood glucose devices, which makes them an interesting source of data to create a model.

The reader captures the data from the sensor when it is closer than 4 cm, and the patient can get their blood glucose values at any moment by reading the data from the sensor. The patient must synchronise the data with the sensor at least once each 8 hours. If not, the sensor erases all the information until the patient does a new synchronisation. The reader can keep blood glucose values for 90 days and provide data through the sensor that is easy to interpret by the patient.

The patients can obtain the data files by connecting the device to a PC. Ten patients provided twelve data files to test the application. The data files were anonymized before their analysis. Some of the data files were incomplete or suffered from a lack of data that had to be considered and managed by the application. The quality of the data files and the results obtained are discussed in section 5.

The data files obtained from a FreeStyle reader contained the following columns:

- ID of the row.
- Date and time that indicates when the record was taken.
- Type of register. The type of registers can take the following values:

0: automatic glucose value register, saved each 15 minutes by the device.
1: manual blood glucose value register, saved in the record after a read by the patient.
2: register of insulin without a numeric value.
3: register of carbohydrates without a numeric value.
4: register of insulin done with a numeric value.
5: register of carbohydrates with a numeric value.
- Blood glucose value in rows with register type 0 (mg/dl).
- Blood glucose value in rows with register type 1 (mg/dl).
- Rapid insulin register without a numeric value in rows with register type 2.
- Carbohydrates without a numeric value in rows with register type 3.
- Units of rapid insulin entered by the patient in rows with register type 4.
- Units of carbohydrates entered by the patient in rows with register type 5.

The carbohydrates and the insulin values (either rapid or slow insulin dose) must be entered by the patient manually before a meal. Other columns such as slow insulin units or data from finger stick were not included in any of the data files analysed for this project, and they were not considered in the data preprocessing procedure.

### 2.2 Decision Trees

Decision tree learning is a supervised learning technique to do classification instances using a series of variables (referred as attributes) [8]. One of the variables must be used as a categorical variable to label the data and the result of the learning process is organised in form of a tree. The purpose of the model is to predict the outcome of the label of a new set of examples by following the rules created by the training examples in the tree.

The implementation of the decision tree classifier used in this project is the one included in the library *scikit-learn*, a package that contains several tools for data mining and data analysis in Python. This classifier is implemented in the class *DecisionTreeClassifier* and can perform multi-class classification on a dataset [9].

Given a matrix *X* of *m* examples with *n* features and a label vector Y, a decision tree splits the instances into subsets recursively such that the examples with the same label are grouped as members of the same class. The impurity in one node of the decision tree is computed using an impurity function, which differs depending on the problem to be solved (either a classification or a regression problem). The measure of impurity used in this application is the Gini index, explained in [10] and [11].

Let t be a node of the decision tree, the Gini index of a node *t* is defined as:

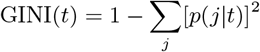

Where p(j|t) is the proportion of observations of class *j* in node *t*.

For example, for a decision tree focused on detecting hypoglycemia patterns like the one in figure 8, the root node contains the following examples:

- Examples classified as “No Hyperglycemia Diagnosis Next Block” 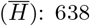: 638.
- Examples classified as “Hyperglycemia Diagnosis Next Block” (H): 1375.

This means that:

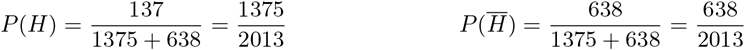

Thus, the Gini index is calculated as follows:

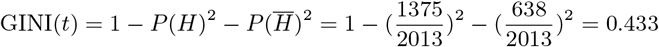

Now, let *t* be a node of the decision tree and *s* split for *t:*

- *P*_*R*_ is the proportion of examples at node *t* that are included in the right child node *t*_*R*_ and *i*(*t*_*R*_) is the impurity of the right child node.
- *P*_*L*_ is the proportion of examples at node *t* that are included in the left child node *t*_*L*_ and *i*(*t*_*L*_) is the impurity of the left child node.

The reduction of Gini impurity (*Gini-Gain*) for a node *t* and a split s is defined as:

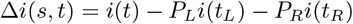

The purpose of the decision tree is to choose the rule in each node that minimise the impurity for that node and repeating the procedure in its child nodes until the maximum depth of the tree is reached. There are several algorithms that try to find the best set of rules to minimise the impurity. The one used by the *DecisionTreeClassifier* is an optimisation of the CART (Classification and Regression Trees) algorithm. CART-based decision trees follow three main steps [11]:

1. Choose the children nodes of each node with less impurity than the parent node.
2. Prune the branches and nodes according to split quality criteria or when the tree is complete.
3. Assign predicted class to the terminal nodes that according to the number examples with greater representation.

The reasons why we have chosen decision trees for this project are the following ones:

- They are easy to understand and interpret, acting like a white box model. The user can find an explanation for each of the rules included in the tree looking at the Boolean logic. This feature fulfils the requirement of using a white box model. Compared to other models such as convolutional neural networks, whose behaviour is difficult to explain to the user, the decisions made by the tree are easily identifiable.
- Decision-tree learners tend to overfit the data if the number of features is very high, but this is not the case of this project, which only uses 22 features for training the model.
- The cost of constructing a decision tree using the efficient implementation of *scikit-learn* and the cost of predict data are *O*(*n*_*features*_*n*_*samples*_*log*(*n*_*samples*_)) and *O*(*log*(*n*_*samples*_)), respectively. The cost-complexity pruning method of CART makes it a fast algorithm for data interpretation, which follows the purpose of the application of being a descriptive tool rather than a predictive tool [12].
- Finding an optimal decision tree is an NP-complete problem and the different implementations use heuristic algorithms to construct them. This flaw is mitigated using ensemble approaches but this option is not considered for this project because they are harder to explain with simple rules. However, finding an optimal tree is not crucial in this application because its main purpose is to find general and repetitive patterns which can be descriptive enough even if the splits are not optimal.

An example code that uses the *DecisionTreeClassifier* can be found in Appendix A. This example code loads one of the data files provided for this study and takes the automatic blood glucose registers (register type 5) to define the following labels: hypoglycemia, hyperglycemia or in range. After defining the labels, the two features used for training the classifier are the prior and subsequent blood glucose values to the current one. These two features are obtained by shifting the column one *±* one period. The purpose of the classifier is to identify the label that corresponds to the current blood glucose value considering only the prior and subsequent blood glucose values, which were taken 15 minutes before/after.

Figure 1 shows the effect of modifying the maximum depth of the tree to control the overfitting. Geometrically, we can observe in the different subplots that the decision trees decompose the attribute space into disjoint subsets using decision rules that are orthogonal to the attribute axes [13]. This rectangular partitioning of the attribute space is a flaw of the decision trees as it does not handle correlated features precisely. Also, pruning the decision tree is necessary, not only because of overfitting control but also because the trees can grow to large dimensions if the number of used features are very high.

**Figure 1:**
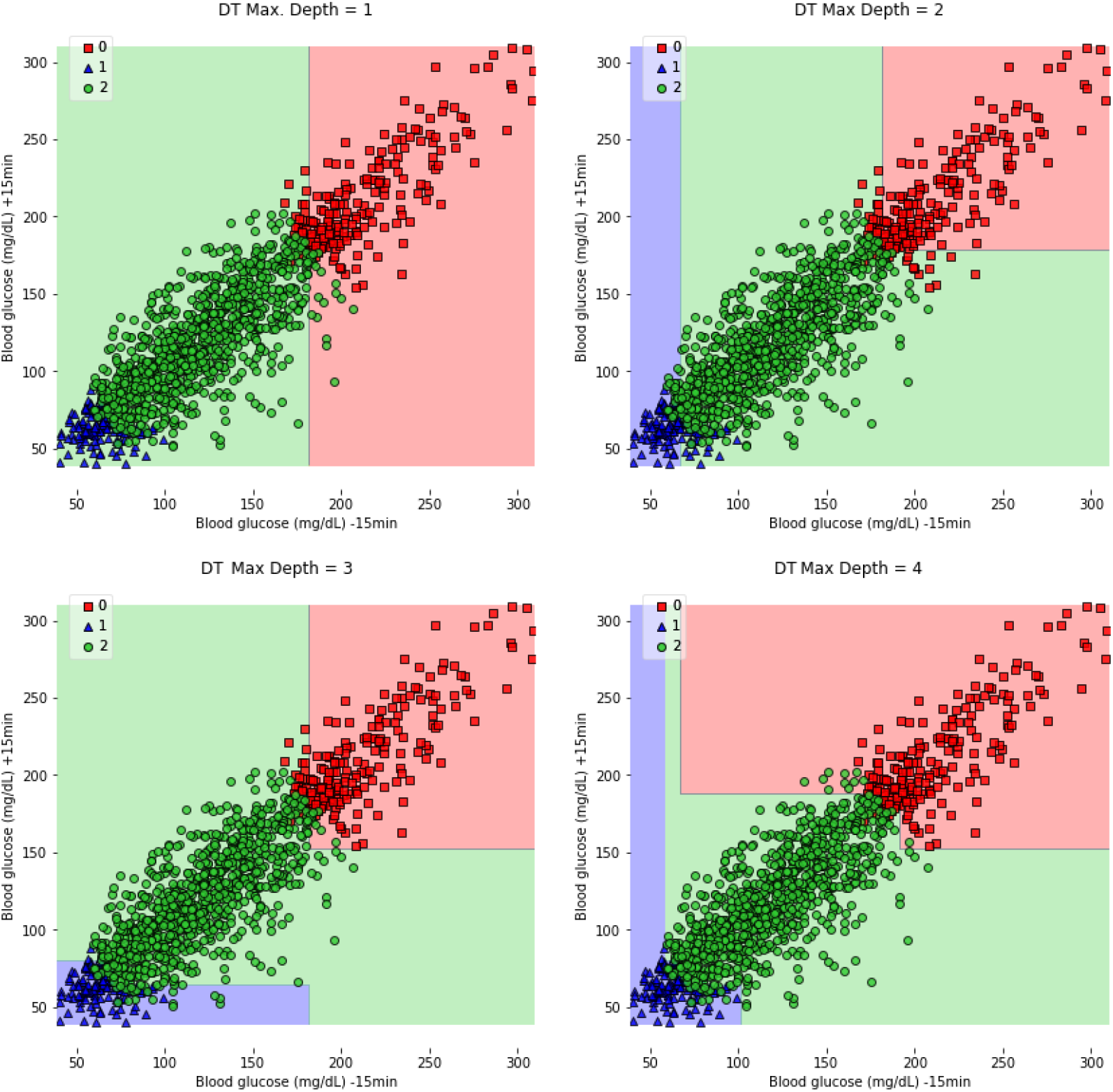
Plot of decision areas in decision trees with maximum depth limit of 1, 2, 3 and 4 levels. The three numbers of the legend stand for hyperglycemia, hypoglycemia and in range situations.

## 3 Design of the application

The development of the application was split into two phases. The purpose of the first step was to develop a self-contained package that could be used both through the Python interpreter or by a web application. The second phase of the application was developing a web application that allow a user to upload a series of data files and obtain the report from a browser. In the two following subsections, we explain the design, the requirements and the technology used for these two modules.

### 3.1 Application core

This main requirement for the application core was to generate a PDF file using a series of data files obtained from a FreeStyle Libre device. The core is purely written in Python, and it uses *pandas* as the main library to upload and manipulate the data.

### 3.2 Web application

The web application has been built using the *Django* framework, written in Python. This framework lets the user define the model of your application and forget about the interaction with the database thanks to its ORM (*Object-Relational Mapping)*. It provides automatic generation of formularies and it has a good interface to build templates. Also, it contains a series of modules that reduces development time and increase the security.

One of the packages included in the web application is the core package of the project, described in the previous subsection. The core is completely integrated into the web application because it does not require a communication protocol since it is also developed in Python. The user needs to fulfil two requirements before starting to use the application:

1. The user must sign up and enter some personal data: name, email and a password. After this step, the user must log in with their account to have access to the service of the application. This authentication protocol has been implemented using the authentication system of *Django*, which handles user accounts and permissions while preserving security.
2. The user must accept a document of terms and conditions of the application. These conditions imply to relinquish some data that it is used by the application to generate the reports. This mechanism has been implemented using the module *Django-termsandconditions*, and it allows to define the terms and conditions from the administration. It also keeps version control of the conditions, forcing a user to accept the terms again if they are updated.

When a logged user accepts the terms and conditions, the application records his IP address and the date of acceptance, giving access to the service.

Through the web interface, the user can upload and delete a series of data files obtained from the FreeStyle devices. It also contains some buttons to select which of the features must be used for training the model and some additional features of the reports like defining the language or including information about the blocks. Figure 2 shows the interface of the page that the user access to generate the report.

**Figure 2:**
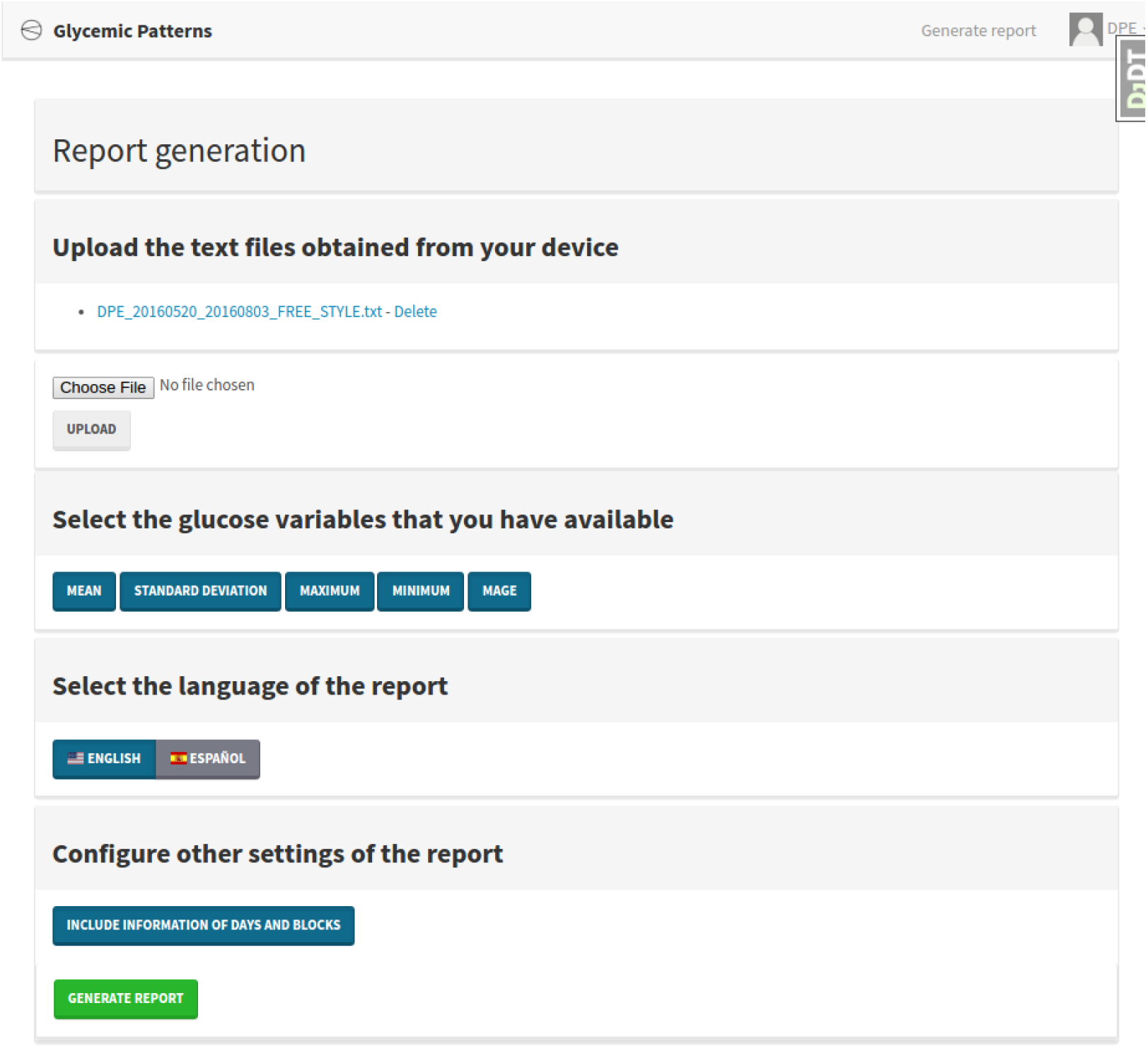
Web interface used by the user to generate a report.

Once the user clicks the button “Generate report”, it sends a POST request to the server. The time necessary to produce the report may vary depending on the resources of the host server and the number of data files that the application must process. After the application core has generated the PDF file, it is returned as a response, and the user can visualise it on the browser or download it to the file system.

## 4 Methodology

### 4.1 Introduction

The application core has been designed to work as an independent component which can be used as a standalone application or as part of an outer application like the web application developed in this work.

The main input of the application core is a series of data files that are used for training the model, and that can be obtained directly from a FreeStyle Libre device. Also, the application provides the possibility of specifying the source language of the data file and some configuration parameters of the decision trees. The output is a report that contains the blood glucose patterns, the decision trees used to extract them and some additional information related to the blood glucose values of the patient. This report can be exported both in PDF and HTML format. In the following subsubsections, we describe the different steps of the program flow (see figure 3).

**Figure 3:**
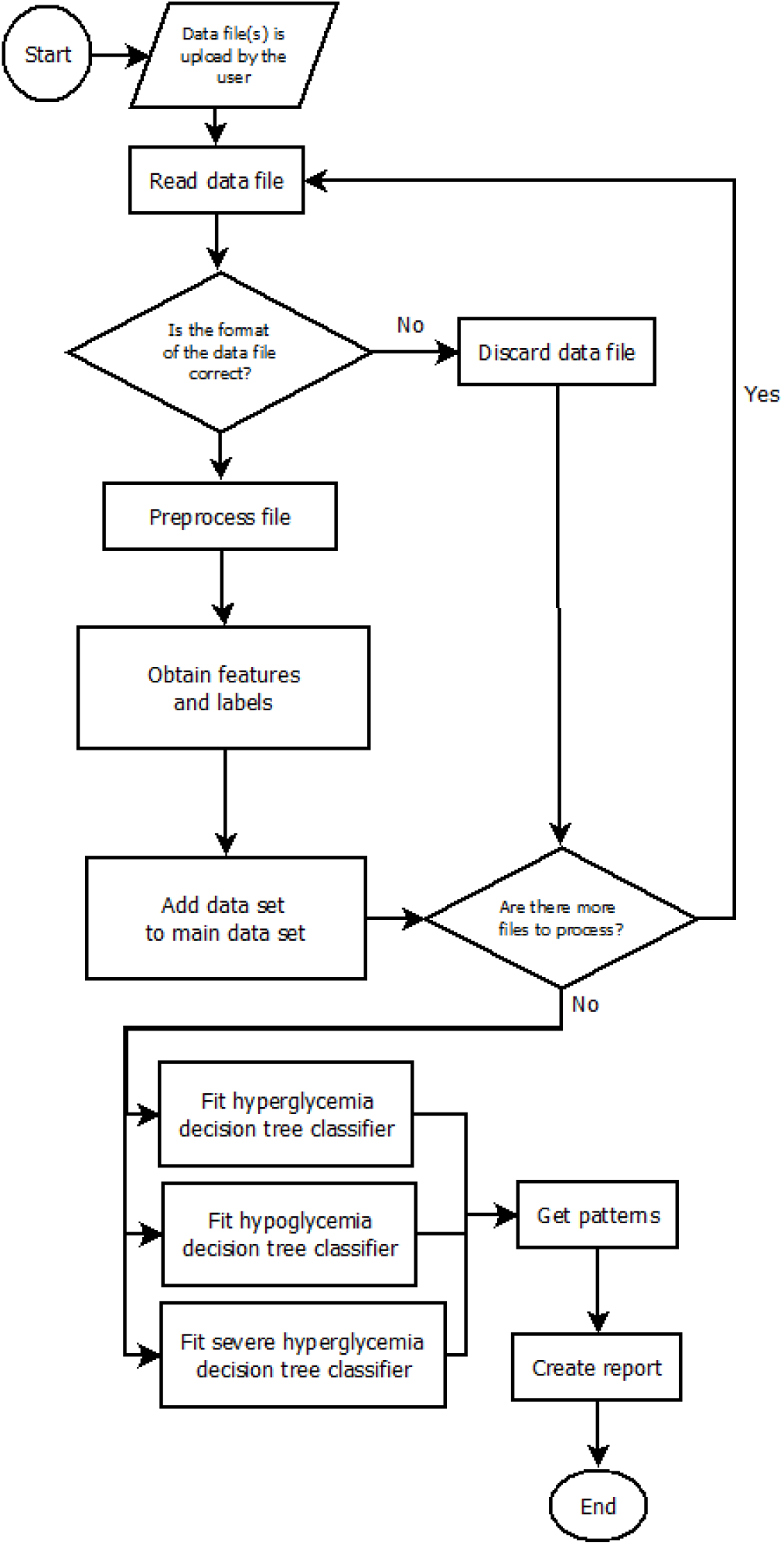
Main flowchart of the application methodology.

**Figure 4:**
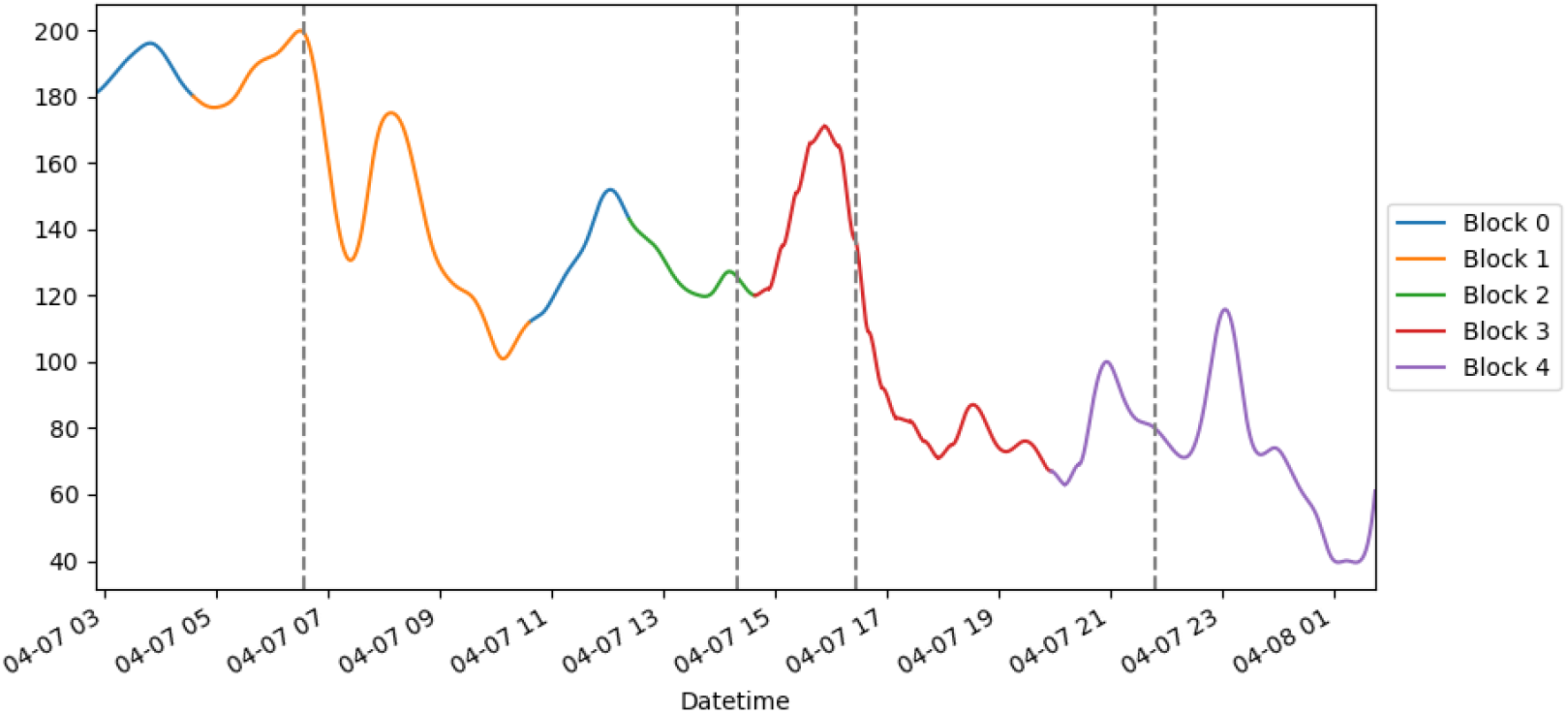
Division in blocks of a sample day. Some blocks are overlapped.

**Figure 5:**
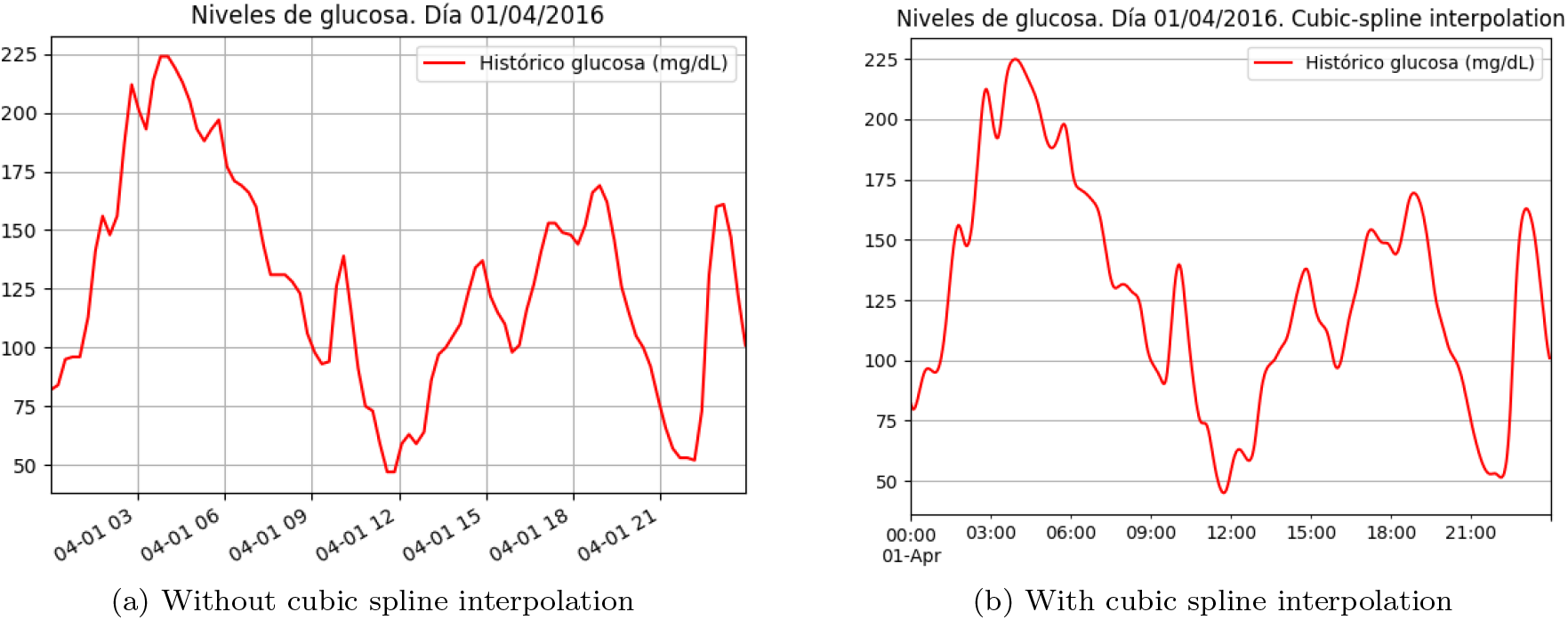
Comparison of plots with/without cubic spline interpolation.

### 4.2 Data preprocessing

The data preprocessing process starts reading a data file (or a set of data files). The *Model* object uses the preprocessing module to deal with the raw data and build the main dataset of the patient. The main dataset is composed of every preprocessed data file and it is used as a single data frame to train the model. This object accepts a list of file paths and the language of the data files’ headers to identify each column. It creates a*Translator* object, in charge of translating the columns to the syntax used inside the code to select the columns.

In the next step, the program iterates all the data files, preprocessing and adding each of them to the main dataset. If the format is not the one expected (different headers, corruption of the data file, wrong extension, etc.), it raises a *DataFormatException*, which can be handled by the web application and reported to the user.

The information in the data files comes from the values recorded by the device, which means that the noise is also encoded in the blood glucose values. The most common anomalies in the data collected by blood CGM systems appear when the device becomes uncalibrated. Although FreeStyle Libre devices do not need to be calibrated, they have an estimation error that can not be corrected with information obtained from any external source (such as finger sticks). Other CGM devices provide the possibility of entering external data to calibrate them, which can be used to detect and fix the anomalies in the recorded values. We made the assumption that the estimation error of the sensor is propagated to the patterns extracted by the model.

The three most important preprocessing techniques used to treat each data file are missing data management, the definition of blocks and smoothing of blood glucose curves.

### 4.3 Dealing with missing data

One of the main issues of the FreeStyle device is that it deletes all the entries in the last 8 hours if the patient do not synchronise the reader with the sensor in that period. As a consequence, the data file may contain some discontinuities in the data. In the application, these gaps are handled dividing one data file in a subset of valid periods. The definition of a valid period is a period of no more than 8 hours without carbohydrate registers. This definition is necessary because some features (like the time elapsed since the last meal) are calculated considering the period between two meals. If this period is too large (greater than 8 hours) because of missing data, it introduces some noise in the dataset that may decrease the accuracy of the model.

Once each period is defined, they are preprocessed separately and incorporated to the main dataset. In any of the following cases, the program discards a period:

- If it has no carbohydrates registers.
- Its duration is less than 24 hours.
- It results in an empty set after preprocessing it.

Another drawback is that the information of meals is only available from one single source: the input made by the patient in the device. If the patient does not specify the number of carbohydrates, it assigns one by default. However, if the patient does not enter any carbohydrate data, this data is not available in the dataset. Some data such as the meal time cannot be inferred easily because a patient typically does not eat at the same time every day. The patient should provide accurate and realistic information so that the report can reveal if carbohydrates have any impact on the patient’s blood glucose values.

If some carbohydrate data are missing, it causes a poor division of the day in blocks and, therefore, decrease the quality of the dataset and the patterns. This issue is notified in the program as a warning (and reflected in the report subsequently) when the mean of carbohydrates of each day in the period is less than one per day.

### 4.4 Define blocks

Each day of the patient is divided into a series of blocks defined by the different meals that the patient has throughout the day. A time window from two hours before to four hours later is defined for each carbohydrate entry (register type number 5), and it only includes automatic measurement of blood glucose values (register type number 1). Each block also contains the number of insulin doses that the patient has taken in the block. The column that contains the number of carbohydrates units and the column without numerical value of carbohydrates units in a meal (one unit) are merged. This intersubsection is also made for the columns that contains the values of rapid insulin doses taken by the patient before a meal.

The next step entails iterating all the days of the dataset and define their corresponding blocks from 0 to *n* (block 0 is the block associated with all entries that have not been included in any other block). For each day, all the occurrences of carbohydrates of that day are used to define its time window and include all the values of carbohydrates and rapid insulin that correspond to that block.

Therefore, several blocks may reference to the same doses of carbohydrates and insulin due to a possible overlapping between blocks. The preprocessor considers this overlapping and it unfolds every overlapped entry in several entries, indicating in one new feature that the block is overlapped.

### 4.5 Smoothing curves

Capillary measurements allow a maximum error of 15 % for glucose levels ≥ 100 mg/dL and *±*15 mg/dL for glucose levels < 100 mg/dL [14]. Even though CGM sensors are useful tools in the management of diabetes due to their high accuracy [15], these errors must be considered by their users. For example, a physician usually smooths the blood glucose values implicitly when he analyses the data.

This technique is used by the program when the visualisation module creates the daily plots included in the report. Cubic spline smoothing is one technique that many physicians identify as the one that is more similar to the implicit smoothing that they apply when they analyse a time series of blood glucose values [5]. Cubic spline smoothing is a cubic spline interpolation that uses a regularisation parameter, and it is described in [16]. We used the implementation that *pandas* library includes, which is a wrapper of the method *interp1d*, present in package *interpolate*, which belongs to the library *scipy*.

### 4.6 Define features and labels

After preprocessing all the valid periods and add them to the main dataset, the process of feature engineering starts. One of the constraints of this project is that the physician and the patient must obtain easily the information of the features used for training the models so that they can understand the patterns and take preventive measures in the future. Many machine learning models are trained using a lot of different features obtained from the time series and complex formulas that increase the predictive power of the model but reduces its comprehensibility drastically.

This application not only uses features that are easy to obtain by the user, but also it allows to select the desired features that will be used to obtain the patterns. Table 2 details a summary of the features that can be used to train the model.

**Table 1:** Unfolding of registers with overlapped blocks.

| Datetime | Glucose auto | Block | Day block | Overlapped block | Carbo block | Rapid insulin block |
| --- | --- | --- | --- | --- | --- | --- |
| 05/04/2016 7:46 | 97 | 1 | 05/04/2016 | FALSE | 1 | 1 |
| 05/04/2016 8:01 | 94 | 1 | 05/04/2016 | FALSE | 1 | 1 |
| 05/04/2016 8:16 | 83 | 1 | 05/04/2016 | FALSE | 1 | 1 |
| 05/04/2016 8:31 | 74 | 1 | 05/04/2016 | TRUE | 1 | 1 |
| 05/04/2016 8:31 | 74 | 2 | 05/04/2016 | TRUE | 1 | 0 |
| 05/04/2016 8:46 | 74 | 1 | 05/04/2016 | TRUE | 1 | 1 |
| 05/04/2016 8:46 | 74 | 2 | 05/04/2016 | TRUE | 1 | 0 |

**Table 2:** Features used in the training process and their descriptions.

| Feature | Description |
| --- | --- |
| $G$ | Value of blood glucose (mg/dL) of the record |
| $G_{D-1}$ | Value of blood glucose (mg/dL) one day before the record |
| $\Delta G_{D-1}$ | Difference between the value of blood glucose one day before the record and the value of blood glucose of the record (mg/dL) |
| $B$ | Block number of the record (0-n) |
| $H$ | Hour (0-23) of the record |
| $OL_B$ | The block of the record is overlapped (True/False) |
| $WD$ | Day of the week of the record (0-6) |
| $\Delta t_{LM}$ | Elapsed minutes since the last meal until the record (0-n) |
| $H_{LM}$ | Hour of the last meal before the record (0-23) |
| $\mu_{B-1}$ | Mean of the blood glucose values of the previous block to the record |
| $\sigma_{B-1}$ | Standard deviation of the blood glucose values of the previous block to the record |
| $max_{B-1}$ | Maximum value of the blood glucose values of the previous block to the record |
| $min_{B-1}$ | Minimum value of the blood glucose values of the previous block to the record |
| $car_{B-1}$ | Carbohydrates (portions) of the previous block to the record |
| $in_{B-1}$ | Rapid-acting insulin (units) of the previous block to the record |
| $\mu_{D-1}$ | Mean of the blood glucose values of the previous day to the record |
| $\sigma_{D-1}$ | Standard deviation of the blood glucose values of the previous day to the record |
| $max_{D-1}$ | Maximum value of the blood glucose values of the previous day to the record |
| $min_{D-1}$ | Minimum value of the blood glucose values of the previous day to the record |
| $car_{D-1}$ | Carbohydrates (portions) of the previous block to the record |
| $in_{D-1}$ | Rapid-acting insulin (units) of the previous block to the record |
| $MAGE_{D-1}$ | Mean Amplitude of Glycemic Excursion of the previous day |

One of the main factors that have a high influence in the appearance of blood glucose disorders is the glycemic variability [17]. One of the main concerns of the type 1 diabetic patients is to control their blood glucose values as much as possible. Despite there is no universally accepted metric to measure glycemic variability, Monnier and Colette state that MAGE (Mean Amplitude of Glycemic Excursion) is an appropriate measure.

MAGE is obtained by calculating the arithmetic mean of all the differences of peak-to-nadir excursions that are greater than the standard deviation of the day. Figure 6 shows one 24-hour period of one sample taken from the FreeStyle device. The module used to obtain the peaks was *peakdetect*.

**Figure 6:**
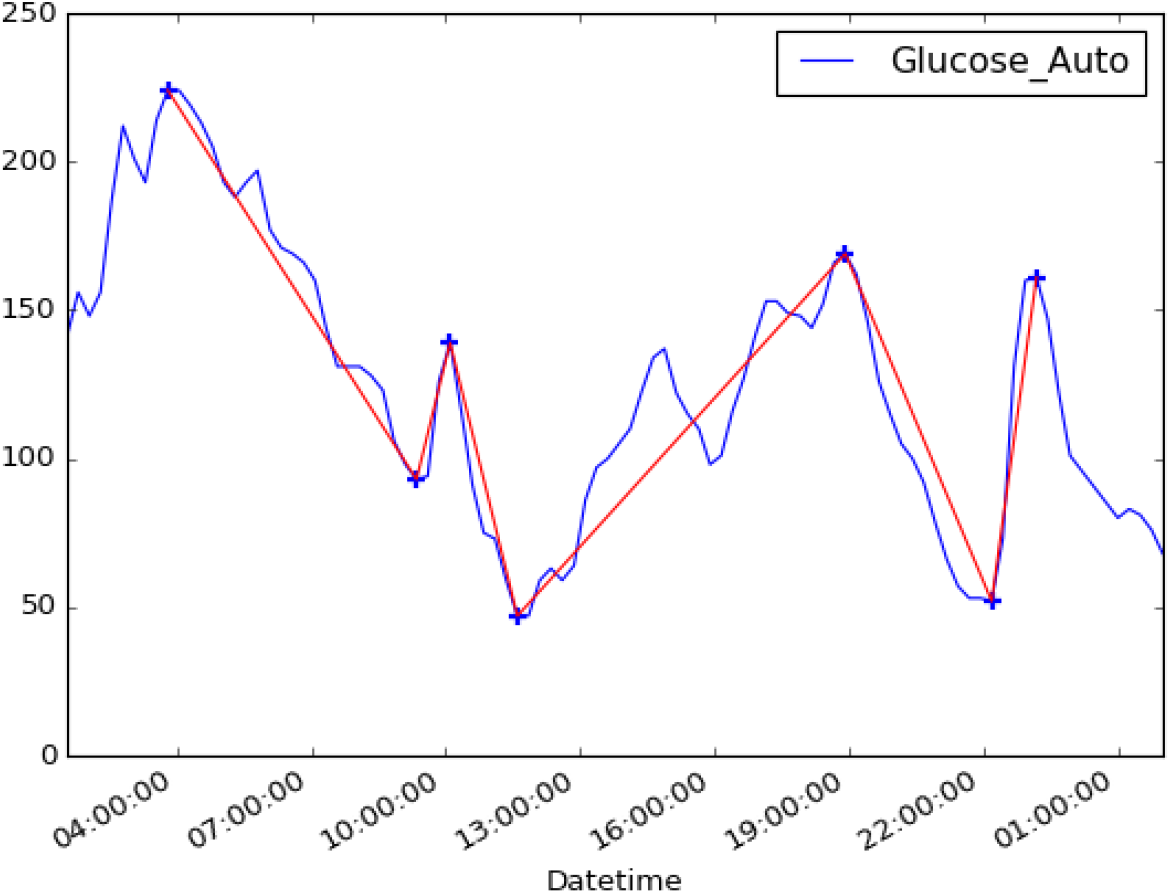
Peak detection and excursions to calculate the MAGE.

The standard deviation of this period is 44.57, and the first excursion that is greater than the standard deviation starts at 4 am. In total, there are six different excursions that are greater than the standard deviation. To obtain the MAGE, we sum them up and divide by 6. The result is the following one:

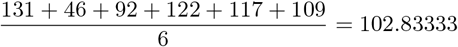

The information that the patient must have at a certain moment was a constraint to define the features. Therefore, the methodology does not include some features that contain information which is derived from periods that finishes after the record time. For example, the number of blocks of each day was not included as a feature because if it is 4 pm and the patient has made three meals until then, may not know how many meals will take until going to sleep. This problem is stressed if the patient varies the number of meals depending on his blood glucose values. Hence, the included features contain information that is available to the patient at any moment such us the blood glucose values of the previous block or the previous day or the day of the week.

Some features need to be adjusted, inferred or removed after the calculation because they are not directly available from the data. One case is *G*_*D*−1_ (value of blood glucose one day before the record), which needs to be rounded to the nearest quarter hour to get *G*_*D*−1_ as the register taken 24 hours before may not exist due to a shifting in the measurement time. After this adjustment, Δ*G*_*D*−1_ can be obtained safety.

The function removes the records with no previous meal, with no data of the previous block or that does not contain values of glucose of the previous day after adding new features to the dataset. These records usually correspond to the first records of the dataset. Once these rows are deleted, the following features are inferred with the mean of the column.

Finally, the function in charge of the feature engineering process defines the labels. Firstly, it adds one column with the diagnosis of the record using the blood glucose value. The four possible diagnoses are the following ones:

1. Hypoglycemia: if *G* < 70 mg/dL.
2. In range: if 70 mg/dL ≤ *G* ≤ 180 mg/dL.
3. Hyperglycemia: if *G* >180 mg/dL.
4. Severe hyperglycemia: if *G* > 240 mg/dL.

It is important to highlight that the hyperglycemia and severe hyperglycemia labels are overlapped when *G* > 240 mg/dL. This overlapping is considered in the next step when the labels are binarized. Binarizing the labels consist in create a binary column for every label and each of these columns is used by a different decision tree in the training process. Once the labels are binarized, the function applies a logical OR to the hyperglycemia column with the severe hyperglycemia columns. One example of this binarization is in table 3.

**Table 3:** Mapping of the diagnosis labels.

| Datetime | G | B | Diagnosis | Hyper-glycemia diagnosis | Hypo-glycemia diagnosis | In range diagnosis | Severe hyper-glycemia diagnosis |
| --- | --- | --- | --- | --- | --- | --- | --- |
| 01/04/2016 20:56 | 78 | 4 | In range | 0 | 0 | 1 | 0 |
| 01/04/2016 21:11 | 66 | 4 | Hypo-glycemia | 0 | 1 | 0 | 0 |
| 02/04/2016 17:50 | 233 | 4 | Hyper-glycemia | 1 | 0 | 0 | 0 |
| 02/04/2016 18:05 | 249 | 4 | Severe hyper-glycemia | 1 | 0 | 0 | 1 |

**Table 4:** Information about the decision trees focused on detecting hyperglycemia patterns.

| Patient | Dates | Number of patterns | Samples (%) of the pattern with more coverage | Impurity of the pattern with more coverage |
| --- | --- | --- | --- | --- |
| Patient 1 | 31/03/16 -14/04/16 | 2 | 10.43 % | 0 |
| Patient 2 | 20/06/16 - 04/07/16 | 0 | - | - |
| Patient 2 | 06/07/16 - 20/07/16 | 2 | 34.24 % | 0 |
| Patient 3 | 07/09/16 - 22/09/16 | 3 | 30.13 % | 0 |
| Patient 4 | 20/05/16 - 03/08/16 | 1 | 67.66 % | 0 |
| Patient 5 | 30/01/16 - 30/06/16 | 3 | 38.76 % | 0 |
| Patient 6 | 04/04/16 - 27/04/16 | 3 | 50.19 % | 0 |
| Patient 6 | 25/05/16 - 29/05/16 | 1 | 71.08 % | 0 |
| Patient 7 | 04/04/16 - 08/04/16 | 0 | - | - |
| Patient 8 | 30/03/16 - 22/06/16 | 3 | 24.40 % | 0 |

**Table 5:** Information about the decision trees focused on detecting hypoglycemia patterns.

| Patient | Dates | Number of patterns | Samples (%) of the pattern with more coverage | Impurity of the pattern with more coverage |
| --- | --- | --- | --- | --- |
| Patient 1 | 31/03/16 - 14/04/16 | 2 | 25.78 % | 0 |
| Patient 2 | 20/06/16 - 04/07/16 | 1 | 63.38 % | 0 |
| Patient 2 | 06/07/16 - 20/07/16 | 2 | 17.52 % | 0 |
| Patient 3 | 07/09/16 - 22/09/16 | 2 | 10.42 % | 0.1728 |
| Patient 4 | 20/05/16 - 03/08/16 | 3 | 29.05 % | 0 |
| Patient 5 | 30/01/16 - 30/06/16 | 3 | 35.49 % | 0 |
| Patient 6 | 04/04/16 - 27/04/16 | 2 | 40.36 % | 0 |
| Patient 6 | 25/05/16 - 29/05/16 | 1 | 44.58 % | 0 |
| Patient 7 | 04/04/16 - 08/04/16 | 1 | 58.59 % | 0 |
| Patient 8 | 30/03/16 - 22/06/16 | 1 | 18.62 % | 0.1653 |

**Table 6:** Information about the decision trees focused on detecting severe hyperglycemia patterns.

| Patient | Dates | Number of patterns | Samples (%) of the pattern with more coverage | Impurity of the pattern with more coverage |
| --- | --- | --- | --- | --- |
| Patient 1 | 31/03/16 - 14/04/16 | 0 | - | - |
| Patient 2 | 20/06/16 - 04/07/16 | 2 | 36.54 % | 0 |
| Patient 2 | 06/07/16 - 20/07/16 | 1 | 19.19 % | 0.2824 |
| Patient 3 | 07/09/16 - 22/09/16 | 1 | 10.47 % | 0.06479 |
| Patient 4 | 20/05/16 - 03/08/16 | 2 | 38.00 % | 0 |
| Patient 5 | 30/01/16 - 30/06/16 | 2 | 19.72 % | 0.1034 |
| Patient 6 | 04/04/16 - 27/04/16 | 2 | 34.85 % | 0 |
| Patient 6 | 25/05/16 - 29/05/16 | 0 | 12.85 % | 0 |
| Patient 7 | 04/04/16 - 08/04/16 | 1 | 82.82 % | 0 |
| Patient 8 | 30/03/16 - 22/06/16 | 0 | - | - |

However, the diagnosis of the current block is not the desired label to extract the patterns because the patient need to anticipate to a risk situation before suffering it. The desired label is one imminent risk situation that the patient can identify and take action to prevent it. This forthcoming risk situation is identified in the next block, which means at least two hours before of the next meal. This way, the patient can pay more attention to his blood glucose values or change his habits for that day to avoid the potential risk.

The function aggregates the data in blocks with a logical OR in such a way that one single register with a positive diagnosis sets a positive diagnosis on the block. Then, the labels are shifted and each row has the label corresponding to the diagnosis of the following block.

### 4.7 Model training

The model is composed of three decision trees, one for each diagnosis and it uses the *DecisionTreeClassifier* class of the *scikit-learn* library [18]. Before training the model, the user has the possibility of specifying via parameters the features to be used in the training process. The classifier has many configurable parameters, some of them necessary to avoid the generation of complex patterns and avoid the overfitting.

The criterion used to make the splits is the Gini impurity rather than entropy in order to minimise misclassification probability. However, the choice of impurity measures has little effect because they are quite consistent with each other [10]. This book also states that the splitting strategy has a greater impact than the impurity measure. The selected strategy to split at each node is to choose the best split rather than a random split. In this case, the best split is the one that minimises the Gini impurity of the children nodes.

Another important parameter that determines the quality of the patterns is the minimum number of samples required to split a node. This parameter is important because it allows stopping splitting those leaves that do not contain a significant number of samples and keep only the relevant patterns. By default, it is set to a 10 % of the total, but this is a parameter that can be configured by the user if he requires patterns with a higher number of samples. It means that all the extracted represent at least the 10 % of the dataset.

The depth of the tree is another parameter that the application allows to configure (by default, is five). This parameter affects the number of rules that compose a pattern. The number of patterns may vary from zero (if the tree has only the root node) to the number of leaves. The algorithm stop splitting when all the leaves have no impurity, when all leaves contain less than the minimum of samples or when the maximum depth is reached. This parameter is quite useful because it allows the user to set the complexity of the patterns and increase their readability and comprehensibility. It also allows to avoid overfitting because very complex patterns may not be general enough to replay a future risk situation.

Other parameters enable to assign different weights to each feature, but this was not desired in this case because all the features are treated equally, having the same possibility of becoming a relevant feature in the pattern. Therefore, the rest of the parameters were set to their default values. Figure 7 shows one decision tree that contains patterns of hyperglycemia.

**Figure 7:**
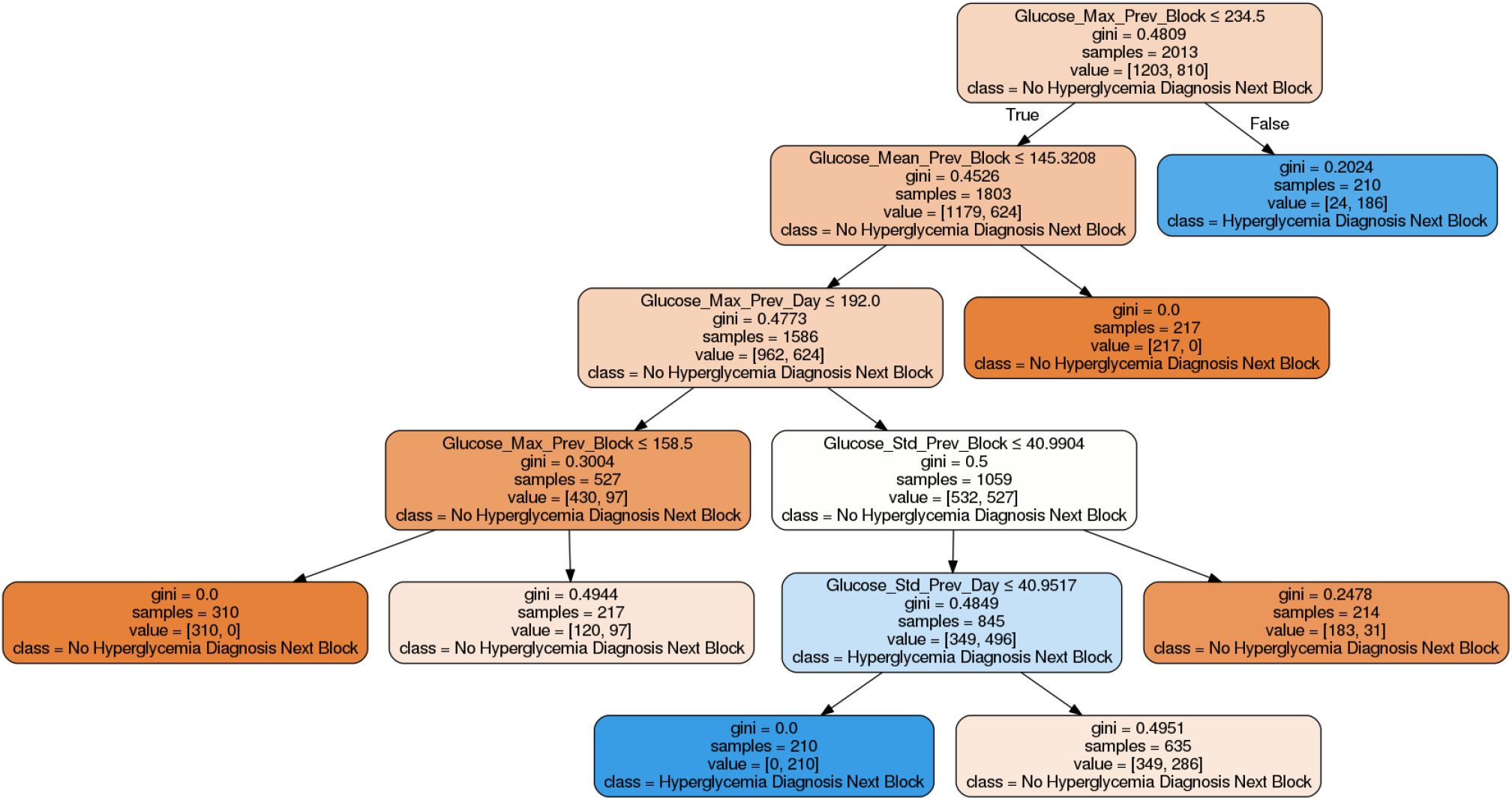
Decision tree that detects patterns of hyperglycemia. Nodes that correspond to risk situations are drawn in blue. Figure 12 shows another example of a hyperglycemia tree.

**Figure 8:**
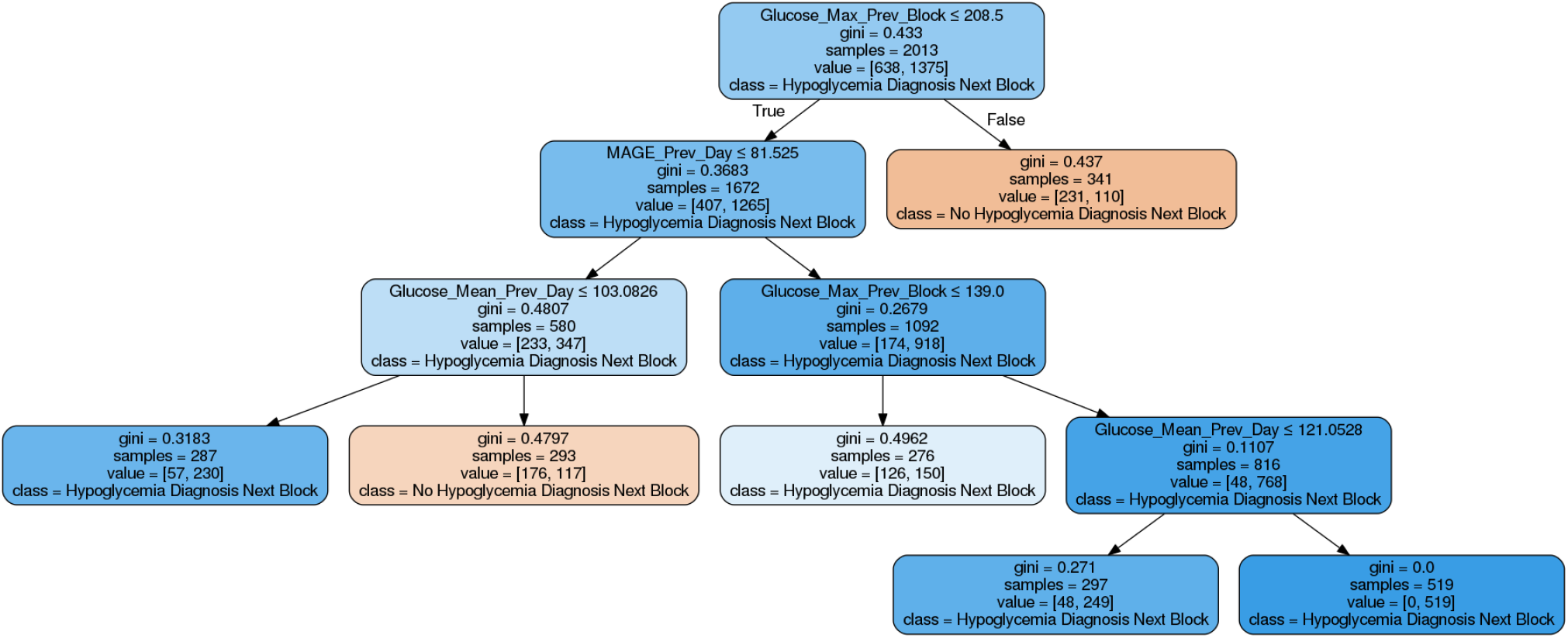
Decision tree that detects patterns of hypoglycemia. Nodes that correspond to risk situations are drawn in blue. Figure 13 shows another example of a hypoglycemia tree.

After fitting the decision trees with the training data and their corresponding labels, the model extracts the patterns from the tree. Recursively, one method traverses all the branches of the tree and keep all of them that has a positive label as a leaf (the class that corresponds to a risk situation). This function allows filtering those branches whose leaf has an impurity value greater than a threshold given by the user. The threshold set by default is 0.3, and if this threshold is reduced, only the purest patterns will be returned by the model, increasing the precision but reducing the recall.

Finally, the model builds a list of *Pattern* objects, composed of *Rule* objects. Each rule is composed of one feature, and operator and a threshold value. For example, a rule defined as *G < 180* means that the current value of glucose must be lower than 180 mg/dL. The model contains an inner class to create compound rules when the same feature appears several times in the pattern, which increases the readability of the pattern. For example, if the rule *G* ≤ 180 is contained in one node and *G >* 135 appears in one of its child nodes, it will be represented as a compound rule that contains one feature, two operators and two thresholds. The compound rule 135 ≤ *G <* 180 is translated as follows: the current value of glucose must be lower than 180 mg/dL and greater or equal than 135 mg/dL.

### 4.8 Report generation

The last step of the methodology is generating the report using the patterns extracted in the previous step together with some additional information obtained from the main dataset. The application uses an HTML template that is the backbone of the report, and the library *Jinja2* renders this template using a dictionary that contains the variables generated in Python. This template is completely dynamic, and its style and content can be customized just like a web page.

The method that generates the report accepts a parameter with the language of the report and configures the *Translator* object according to this parameter. Therefore, all the variables that the method passes to the template are previously translated, and the help page that is included at the end of the report also changes depending on the language. The method can generate the report in both PDF and HTML format. The images that appear the report (such as the tree graphs or the plots) are saved previously in a folder with a unique identifier provided as a parameter in case the user application wants to use them for other purposes.

The report is structured in four parts:

1. Information about the patterns along with their rules for each risk situation.
2. Decision trees that have been used to obtained the patterns.
3. Statistics and division in blocks of every day in the dataset.
4. Information about the contents of the report.

The warnings issued by the application core are displayed at the beginning of the report. For example, if the quality of the dataset is not sufficient, a message alerts the patient that the patterns may not be accurate. After these warnings, the first part contains the patterns of hyperglycemia, hypoglycemia and severe hyperglycemia. If it has not been possible to obtain patterns for any of these conditions, the report does not include the subsection of patterns for that condition. The following elements compose each pattern:

1. Set of rules that describe the risk situation in the next block.
2. Number of samples of the dataset that are obtained by the defined rules in the pattern together with the percentage of samples regarding the total of the dataset.
3. Impurity of the pattern.
4. Positive samples classified as a risk situation in the next block along with the percentage of samples regarding the total of positive samples.
5. Negative samples classified as a situation without risk in the next block along with the percentage of samples regarding the total of negative samples.

The second part of the report shows the graphical representation of each tree. These images provide additional information that cannot be obtained from the patterns. The trees show all the patterns, including the ones that do not define a risk situation. The colour scale of each node determines both the majority class and the impurity. The colours of the tree are a good indicator to identify if the patient tends to suffer from a condition more than another. For example, in figure 8, it can be observed that the patient tends to suffer from hypoglycemia at least one time in each block.

The intensity of the colour increases as the impurity of the node decreases. It means that the patterns that end in a leaf with an intense colour (either blue or orange) are the ones that are capable of separate samples that only belongs to one class. Also, each node also indicates its impurity value, the number of samples that follow its rule, the number of samples that belongs to each class and the majority class (which determines the label of the node).

The third part of the report is optional and appears in the report only if the user passes a certain parameter. This information can help the patient or physician to see where the decisions of the tree come from and see the blocks that usually lead to a risk situation. It includes statistics per block and day such as the maximum and minimum, mean and standard deviation of the blood glucose values and the glycemic variability measured with MAGE.

Finally, the fourth part is just information that helps the patient and the physician to understand the concepts that are detailed in this document and makes each report a self-contained piece of information. In Appendix B, there is a sample of a generated report containing all the patterns, the information of the blocks of one day and the information about the report.

## 5 Experimental results

Ten patients provided twelve different data files for developing and testing the application. The mean age of the patients was 34 years with a standard deviation of 10.34 years. Regarding their height, the patients had an average height of 165 cm with a standard deviation of 9.12 cm. Finally, their weights values had a mean of 66 kg with a standard deviation of 16.75 kg.

Two of the data files provided by these patients did not contain any register of carbohydrates (register type 5), and the application returned an error because no block could be defined. The ten remaining files contained a broad range of rows from 427 to 11444 and a mean of 2668 rows. There is no perfect positive correlation between the number of days and the number of rows because some of the data files contained time gaps. These time gaps were produced if the patient did not synchronise the sensor with the device at any moment in a period of 8 hours (as it was described in subsection 4.3). These gaps also vary in a wide range of time from 8 hours to 14 days. The minimum period is 4 days; the maximum is 152 days, and the mean is 40 days.

Seven out of ten files produced a warning by the application, which reported that they had an average of less than three registers of carbohydrates per day. This warning may be considered as an indicator of the quality of the data files and, consequently, the quality of the generated patterns. Some of the features calculated from the data rely on the carbohydrates information entered by the patient such as the blocks or the minutes since the last meal. If this information is incomplete or faulty, the generated patterns also are likely to be imprecise. The patients that provided a high-quality data files were patient 1, patient 3 and patient 6 (the data file corresponding to the period 25/05/16 - 29/05/16).

The decision trees were analysed considering several parameters:

1. Number of patterns extracted.
2. Percentage of samples regarding the total, and impurity of the pattern with the largest coverage. The coverage was defined as the proportion of samples that fulfilled the rules of the pattern.
3. AUC score of the decision tree using 5-fold cross-validation.
4. The degree of importance of its features.

AUC (Area Under Curve) measures the overall performance of a model and its values usually fall in a range from 0.5 to 1. The minimum value is obtained by a model that makes random predictions and the maximum value is obtained by a perfect classifier. Values lower than this range indicates that the model performs worse than a random classifier [19]. AUC metric is derived from the ROC (Receiver Operating Characteristic) plot and it is the most common quantitative index that describe it [20]. This curve shows the relation between the true positive rate and the false positive rate as its discrimination threshold varies. *K*-fold cross-validation splits the dataset into *k* subsets (folds) of approximately equal size. After splitting the dataset, the model is trained *k* times rotating *k-*1 folds and validating over the remaining fold [21]. This evaluation was done using the AUC metric and rotating over five folds of each dataset for this project. An existing implementation of cross-validation is provided by the method *cross val score* included in the package *model selection* of the *sklearn* library.

The importance of the features of each tree is calculated using the Gini importance. [22]. The Gini importance of a feature is computed as a normalised total reduction of the Gini index in each split done considering the feature. The higher the Gini importance is, the more informative is a feature. After computing the importance of each feature in each tree for all the datasets, the results were grouped by the mean of the importance of the features for each type of tree. Figure 9 shows a summary of the importance of all the features.

**Figure 9:**
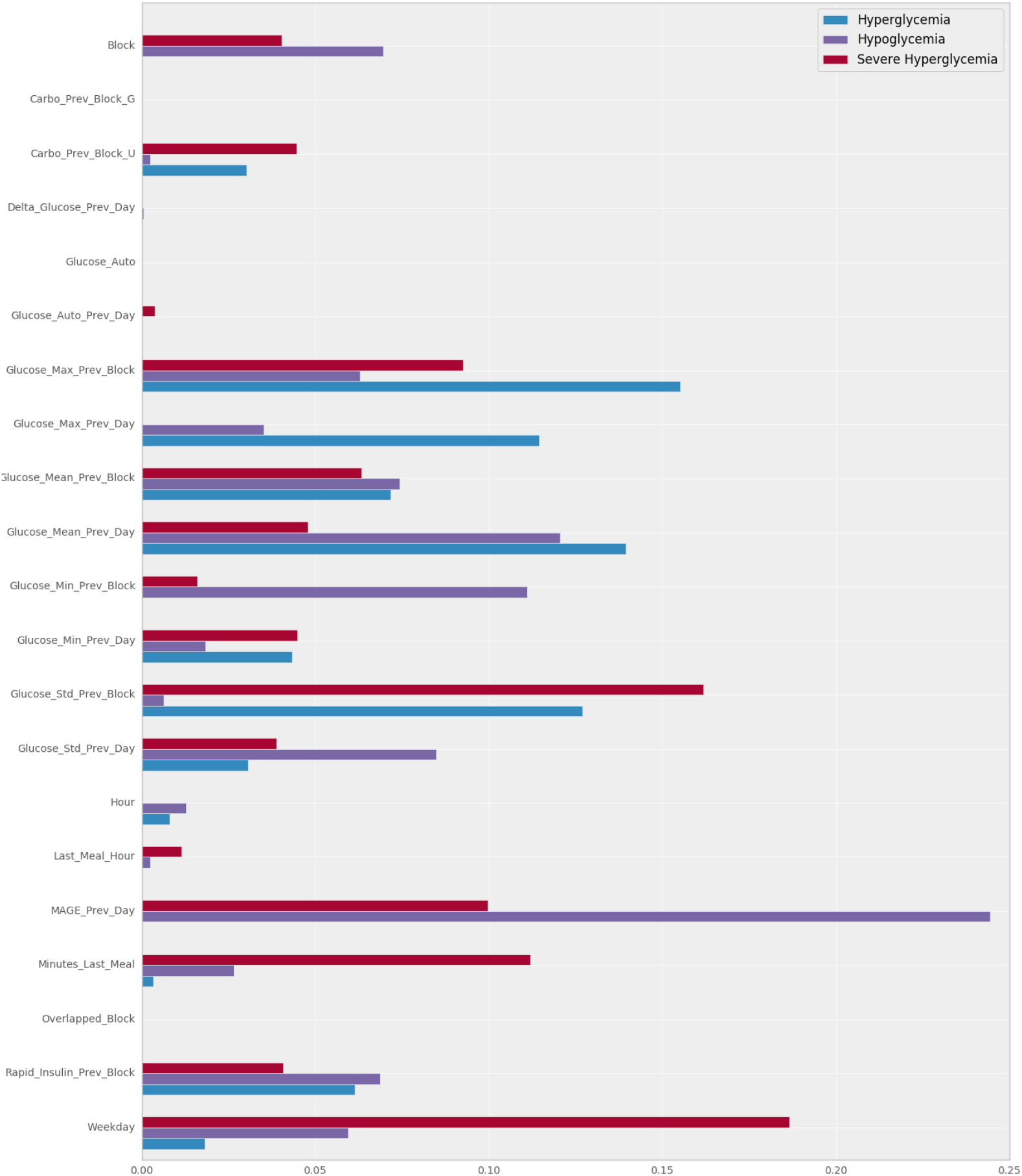
Average Gini importance of each feature for the three types of decision trees, obtained after processing ten different datasets.

All the decision trees that detected patterns of hyperglycemia provided at least one pattern with zero impurity. The largest patterns had a coverage average of 40.86 % and detected a mean of 1.8 patterns for all the data files. Two of the ten hyperglycemia decision trees did not provide any pattern, and the ones that provided only one pattern, it had a high coverage (71.08 and 67.66 %). The mean AUC score of the eight trees that produced at least one pattern was 0.6703, with a maximum of 1 and a minimum of 0.4186. It is relevant to highlight that the tree that scored an AUC of 1 was obtained using a data file that contained only 427 rows and a mean of more than three carbohydrates entries per day.

The most relevant feature of the hyperglycemia decision trees was *max*_*B*−1_, with an average value of 0.1552. With similar values, there were other features related the blood glucose values of the patient such as *µ*_*D*−1_ and *σ*_*B*−1_ with average values of 0.1395 and 0.1270. The following set of features were not used in any of the trees to describe a pattern:

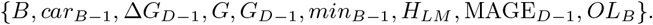

Regarding the hypoglycemia trees, all of them produced at least one pattern. The maximum coverage of a pattern was 63.38 %, with no impurity. The average of detected patterns 1.8, just as hyperglycemia decision trees. The mean AUC score of the trees was 0.6197, lower than the hyperglycemia trees with a maximum score of 1 and a minimum of 0.2834. The tree that scored the maximum score also was produced by a file with a low number of rows (407).

The most relevant feature of the hypoglycemia decision trees was MAGE_*D*−1_, with a value of 0.2443. The second and third most importance features were *µ*_*D*−1_ and *min*_*B*−1_, with average values of 0.1206 and 0.1110. It is relevant the difference between the primary feature and the second most important: the value of the first one is twice as high as the second one. MAGE_*D*−1_ also had the highest value of Gini importance of all the trees, which means that is the most informative feature to describe a pattern (of hyperglycemia in this case). The set of the following features had an average Gini importance of 0: {*car*_*B*−1_, *G, G*_*D*−1_, *OL*_*B*_}.

The decision trees focused on detecting severe hyperglycemia patterns recognized patterns in seven cases, with a mean of 1.1 patterns, lower than the hyperglycemia and hypoglycemia decision trees. The mean AUC score was close to the hyperglycemia decision trees score (0.6774), with a maximum of 1 and a minimum of 0.4463. The decision tree that scored the maximum AUC used the same data file that the decision tree of hyperglycemia that scored the maximum AUC, which contained the lowest number of rows of all the data files. The average coverage of the patterns was 31.81 %, lower than the other two types of decision trees.

The most informative feature of the patterns of severe hyperglycemia was *WD*, with a value of 0.1865. The second and third features in the ranking were *σ*_*B*−1_ and Δ*t*_*LM*_ with an average Gini importance of 0.1619 and 0.1120. The set of features that were not considered in any of the patterns is the following one:

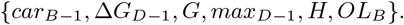

## Results discussion

The coverage results indicated that the patients of the study either were prone to suffer further from hypoglycemia than hyperglycemia or that the hypoglycemia patterns can be described better by the features used by the decision trees. The first theory may follow the intuition that the diabetic patients that decided to participate in this study has a better control of their blood glucose values and therefore, a tendency to keep them low.

As it was expected, the coverage of the severe hyperglycemia patterns was lower than the ones produced by the hyperglycemia decision trees. This phenomenon happened because the number of positive cases of severe hyperglycemia was a subset of the number of positive labels of hyperglycemia and consequently, their coverage should be lower or equal than the largest hyperglycemia pattern.

Furthermore, the results indicated a negative correlation between the number of rows and the AUC score. This negative correlation is more marked in severe hyperglycemia decision trees (−38.51) and can be appreciated in figure 10. A possible explanation for this result may be that the data files with a low number of rows are biased due to their size and the patterns are easier to model than the ones with a higher sample size. Some studies like [23] showed that the AUC score of a single dataset tends to increase when the sample size is larger.

**Figure 10:**
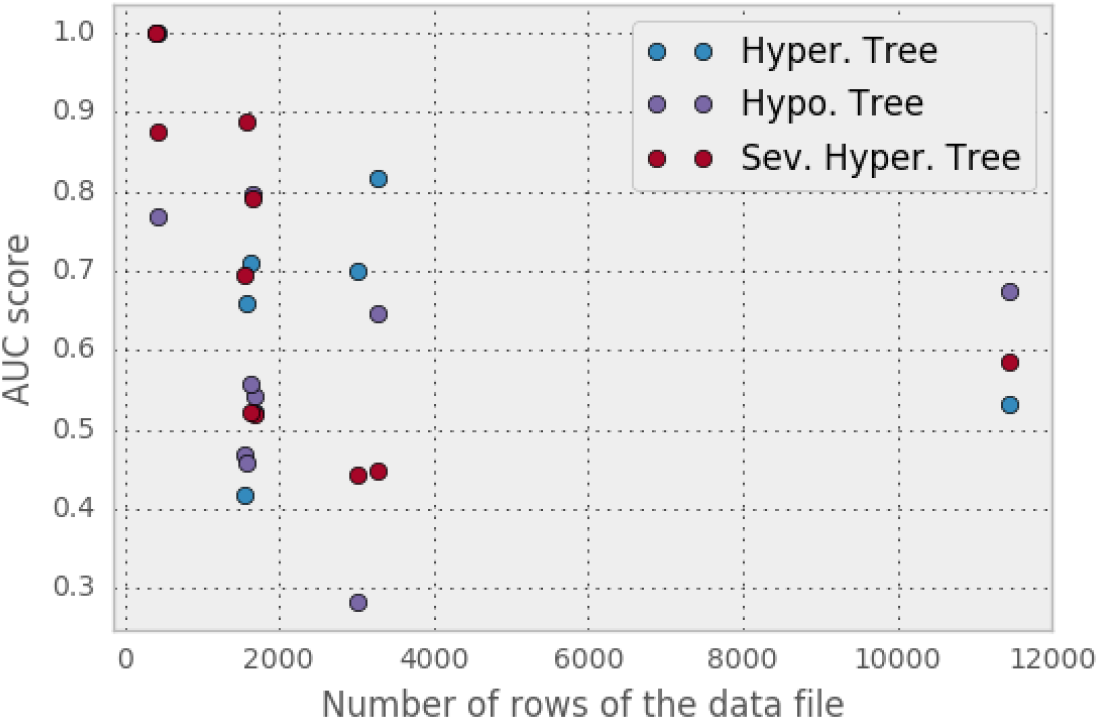
AUC score of the different decision trees and the number of rows of the provided data file.

**Figure 11:**
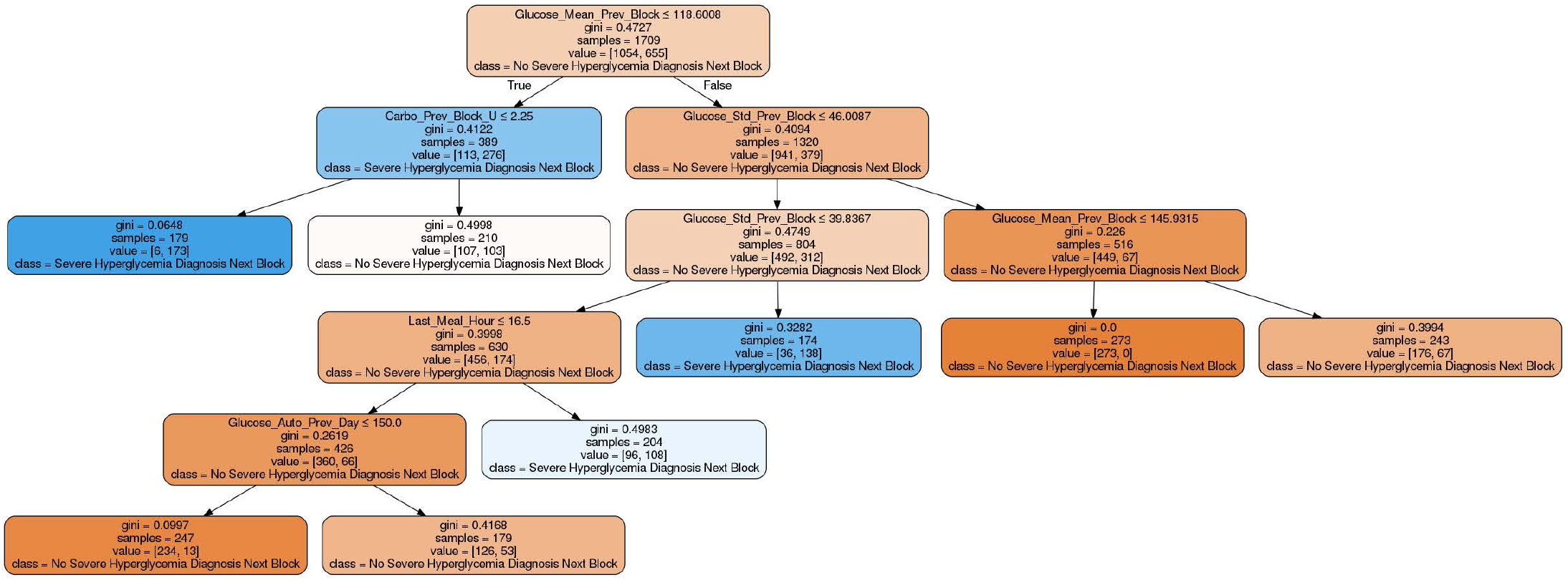
Decision tree specialised in detecting patterns of severe hyperglycemia.

**Figure 12:**
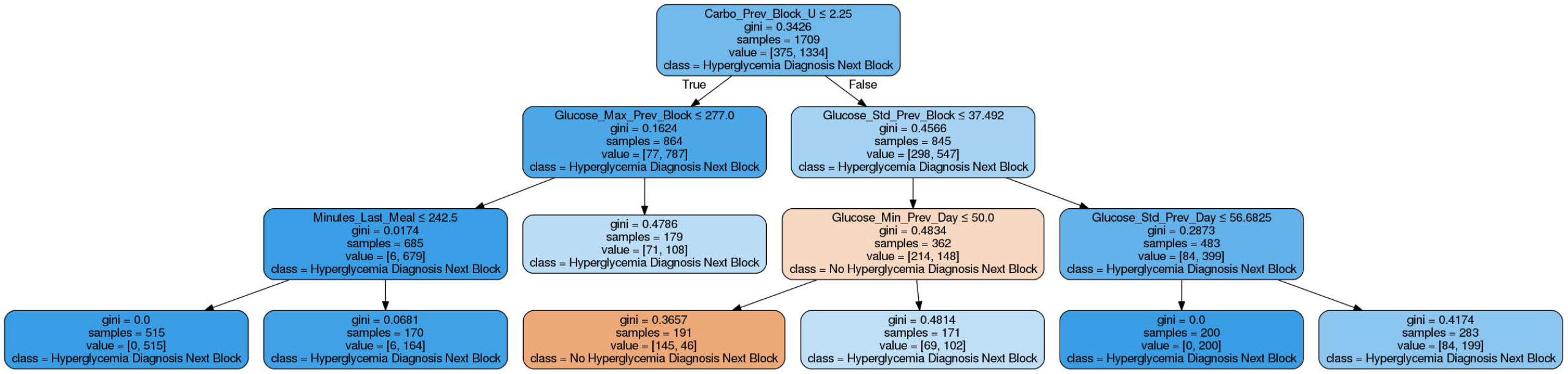
Decision tree specialised in detecting patterns of hyperglycemia.

**Figure 13:**
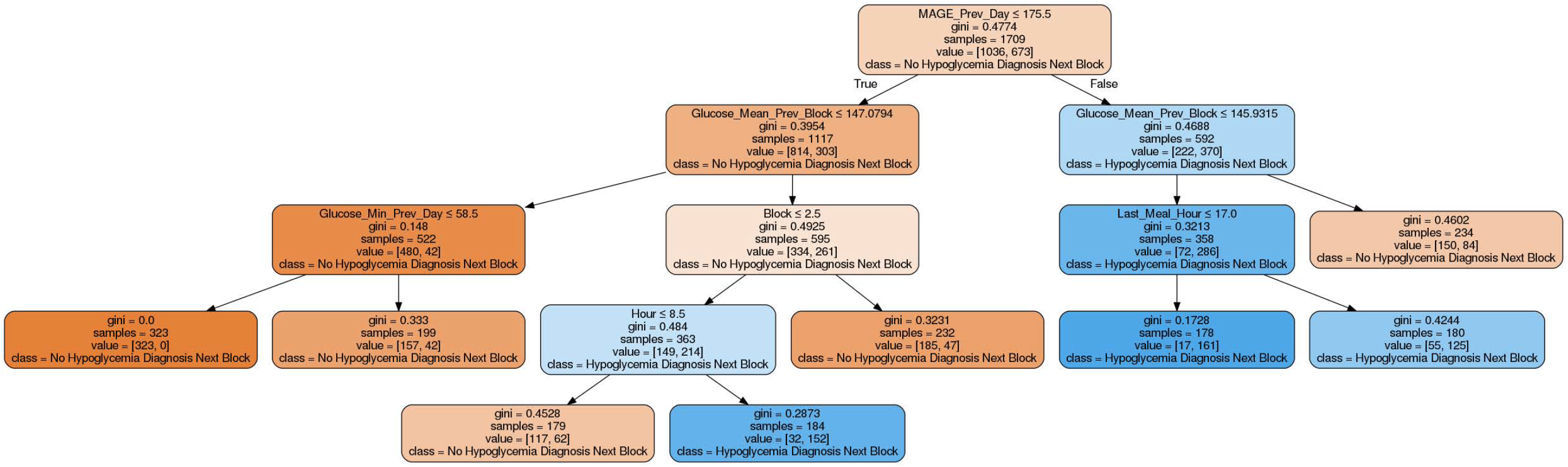
Decision tree specialised in detecting patterns of hypoglycemia.

However, in this case, we are comparing the score of decision trees that use datasets of different patients. Thus, the bias produced by the sample size and the quality of the carbohydrates entries has a high impact on the AUC score of the trees. It was no possible to establish a fair comparison between the AUC value of the decision trees of each patient because some variables such as the size of the dataset, the number of carbohydrates entries and the number of gaps have a high variance in each of them. This negative correlation may not be a relevant result as the data files were not taken under the same circumstances and the data quality differs in each of them.

The analysis of the importance of the features led to several interesting conclusions. The first important result is the fact that hypoglycemia patterns used the glycemic variability of the previous day (MAGE) as the most informative feature. This phenomenon also appears in [24], which states that MAGE was the only measure of glycemic variability significantly higher in the group of patients with repeated hypoglycemia.

The average Gini importance of MAGE_*D*−1_ in hypoglycemia trees shows a contrast with the value obtained for hyperglycemia trees, where it was not used to do any split, resulting in a Gini importance of 0. This result has multiple possible interpretations, and it is an interesting topic to discuss. The hypothesis about this phenomenon is that patients who suffered a high glycemic variability the previous day tend to over-control their blood glucose values the next day, leading to hypoglycemia situations.

Another important result is that it does not exist a strong correlation between the importance of the features of hyperglycemia trees and severe hyperglycemia trees. Whereas the features that provide more information to the hyperglycemia trees were statistics related to blood glucose values, the decision trees focused on severe hyperglycemia used more the variables of time such as the day of the week or the minutes elapsed since the last meal to do their splits. This result may indicate that severe hyperglycemia situations can be more predictable by looking into the variables related to the lifestyle of the patient or the actions that are out of its routine. The decision trees focused on detecting hyperglycemia situations should use blood glucose value to detect the risk situations rather than variables of time.

Lastly, some features were expected to be important because the provide values highly correlated with risk situations. For example, a patient who had a very low minimum of blood glucose the last block is more likely to suffer from hypoglycemia at any moment of the next blocks than a patient with a peak of blood glucose in the last meal. However, other features did not provide any information to the decision trees. These features were the current value of glucose, the difference of blood glucose with the previous day and the present value of blood glucose. This lack of importance suggests that considering one single value of blood glucose do not provide information gain to the trees and it is necessary to consider statistics and indicators that apply to a series of blood glucose values to increase their predictive power.

## 6 Conclusions

An application that applies machine learning techniques in its back-end sometimes suffers from a lack of transparency as the difficulty to track the origin of the provided results. A rigorous assessment of the medical advice given by these algorithms is essential to promoting the development of these applications and their use by patients and physicians [25]. Some questions about the use of black box algorithms in decision-making support systems must be considered before they start to work in medical environments. The main feature of the application developed in this project is to be transparent to the user. The patient or physician can know about the origin of the patterns and use their criteria to either consider them valid or reject them.

The web application together with the core is ready to be tested with real data and be deployed to a production environment after considering some security issues that involves data protection and resilience to cyber attacks. Despite the limitations, all the requirements defined at the beginning of the document has been fulfilled, and the application can obtain some patterns that are potentially significant if the quality of the data source is high.

This work has revealed us the potential of using and process the available data to raise the living standards of people that suffer from health conditions.

Carrying out a data science experiment is challenging because it is hard to estimate if the results are going to be useful or significant at early stages of the development process. This risk is inherent to a data-driven approach because the data do not always contains all the information needed for offer an appropriate service to the end user. This methodology seemed to worked in this project as the final application provides a service to the user, placing the data at the centre of the application. However, many improvements can be made in the application, which are discussed below.

At the beginning of the project, we wanted to tackle the problem with an unsupervised learning technique and the use of clustering to detect the patterns of glycemia. However, this technique was reconsidered as a supervised learning task to try to predict a future risk situation in the blood glucose values of the patient. The idea of dividing each day of the patient in blocks, defined by the ingestion of carbohydrates, can be used to generate new features and develop an online predictive tool in future projects.

One limitation of the application is that the patterns are not extracted in real time, but the patient needs to upload the file to a web application to obtain the results. This limitation is also present in the FreeStyle Libre application, which requires a connection from the reader to a computer through a USB cable. This project has suggested a way of detecting patterns, but the algorithm and the features can be modified to turn the model into a powerful predictive tool in real time. There are several machine learning techniques based on decision trees that use ensemble methods to obtain a better predictive performance.

One example of an ensemble classifier is random decision forest, which uses a multitude of decision trees, capable of mitigating the distortion caused by the existence of noise in the dataset or reduce the overfitting of the model. Another example of an ensemble technique is the gradient tree boosting, which makes use of decision trees as base learners and distribute the weights to identify the trees capable of identifying the most intricate patterns. These ensemble models have a better performance that a single decision tree but it reduces the transparency of the model. Thus, they broke the constraint of this project of creating a transparent model, and they were not used to detect the patterns.

Another improvement can be done in the feature engineering process. The application only makes use of the features that are available to the physician or the patient and give freedom of choosing which features are the ones he wants to use to discover new patterns. If the purpose of the model is to predict a future risk situation by using all the features it has at its disposal, tens of features can be extracted from time series. Also, many libraries are capable of making an automatic extraction of features from a time series. An example of this library is *tsfresh*, whose algorithm is described in [26]. This library extracts up to 100 features from a time series in parallel and makes feature selection of the most relevant ones.

Some parts of the code, especially the generation of the report, can speed-up by parallelizing its execution. Python has many libraries to make parallel computing like *dask*. This library allows optimising an application using dynamic task scheduling and multicore execution. The application could offer a better experience to the patient if the time to generate the report is reduced and this is an excellent opportunity for improvement.

Finally, using the features of the dataset, it is possible to define alternatives labels to discover new patterns. Two examples of possible phenomenons that can be analysed using the data obtained from FreeStyle Libre are the Dawn phenomenon and the Somogyi effect (this last one is well described in [27]). The Somogyi effect consists on a rise of the blood glucose levels as a reaction of the body to a situation of nocturnal hypoglycemia, resulting in high blood sugar levels in the morning. The model could be adapted to detect if a patient suffers from this effect and report it so that the physician can provide with some advice to avoid it.

## Data Availability

Data of the patients is not publicly available. It could be available on demand and after signing a Non Discluser Agreement.

## Acknowledgements

This work has been funded by:

- Fundación Eugenio Rodríguez Pascual 2019-2020.
- Ministerio de Economía y Competitividad under grant TIN2014-54806-R.
- Ministerio de Ciencia, Innovacion y Universidades under grant RTI2018-095180-B-I00.
- Comunidad de Madrid under grant B2017/BMD3773 (GenObIA-CM).

## A *DecisionTreeClassifier* example

The following code written in Python shows one example of how to use the libraries *sklearn* and *mlxtend* to train four decision trees and display the decision areas of the trees. This code has the purpose of studying the effect of varying the maximum depth constraint in the decision areas of each decision tree. The decision trees are trained to predict hyperglycemia and hypoglycemia situations using blood glucose values of the previous and subsequent registers. The resulting plots can be seen in figure 1.

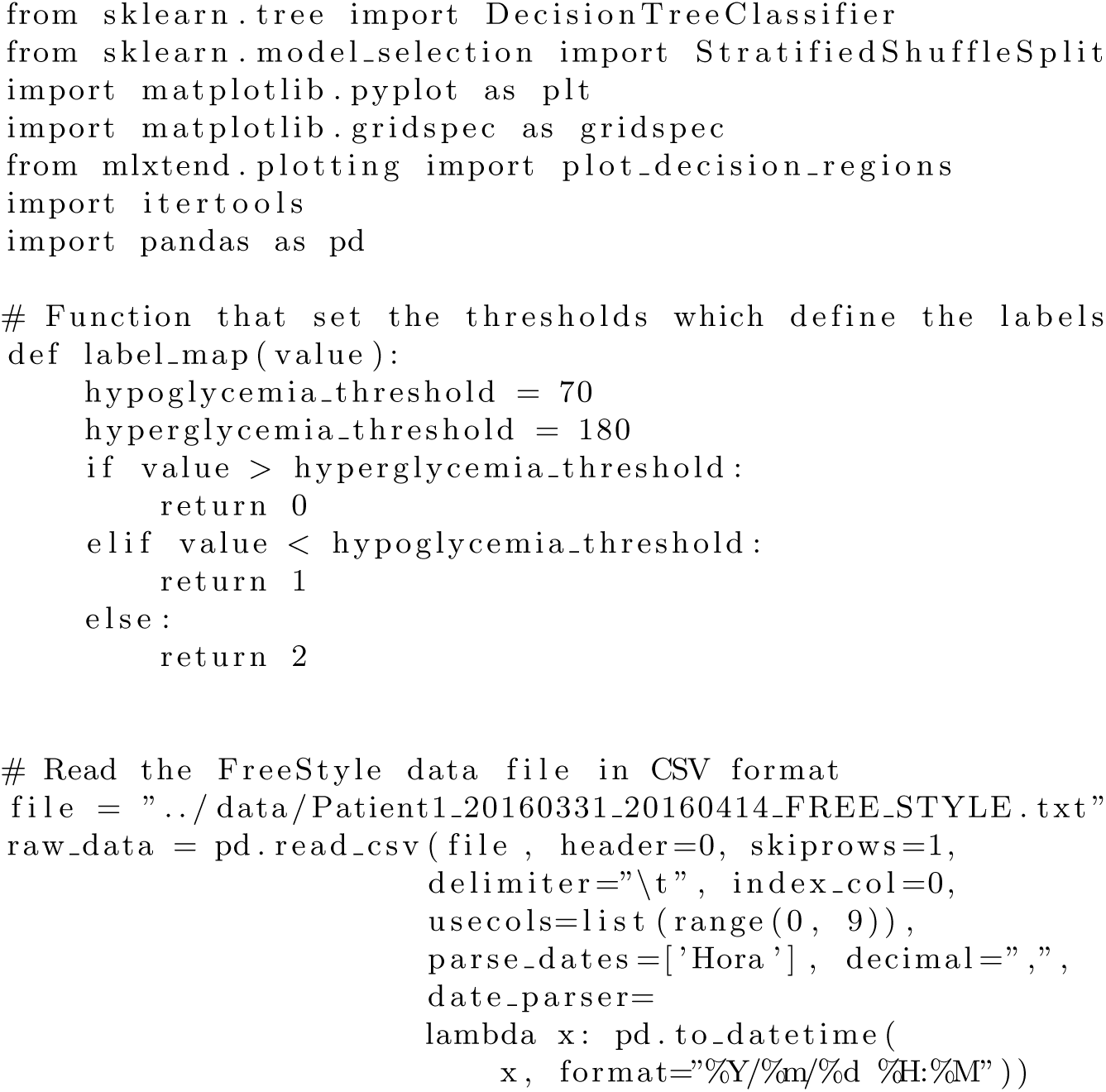
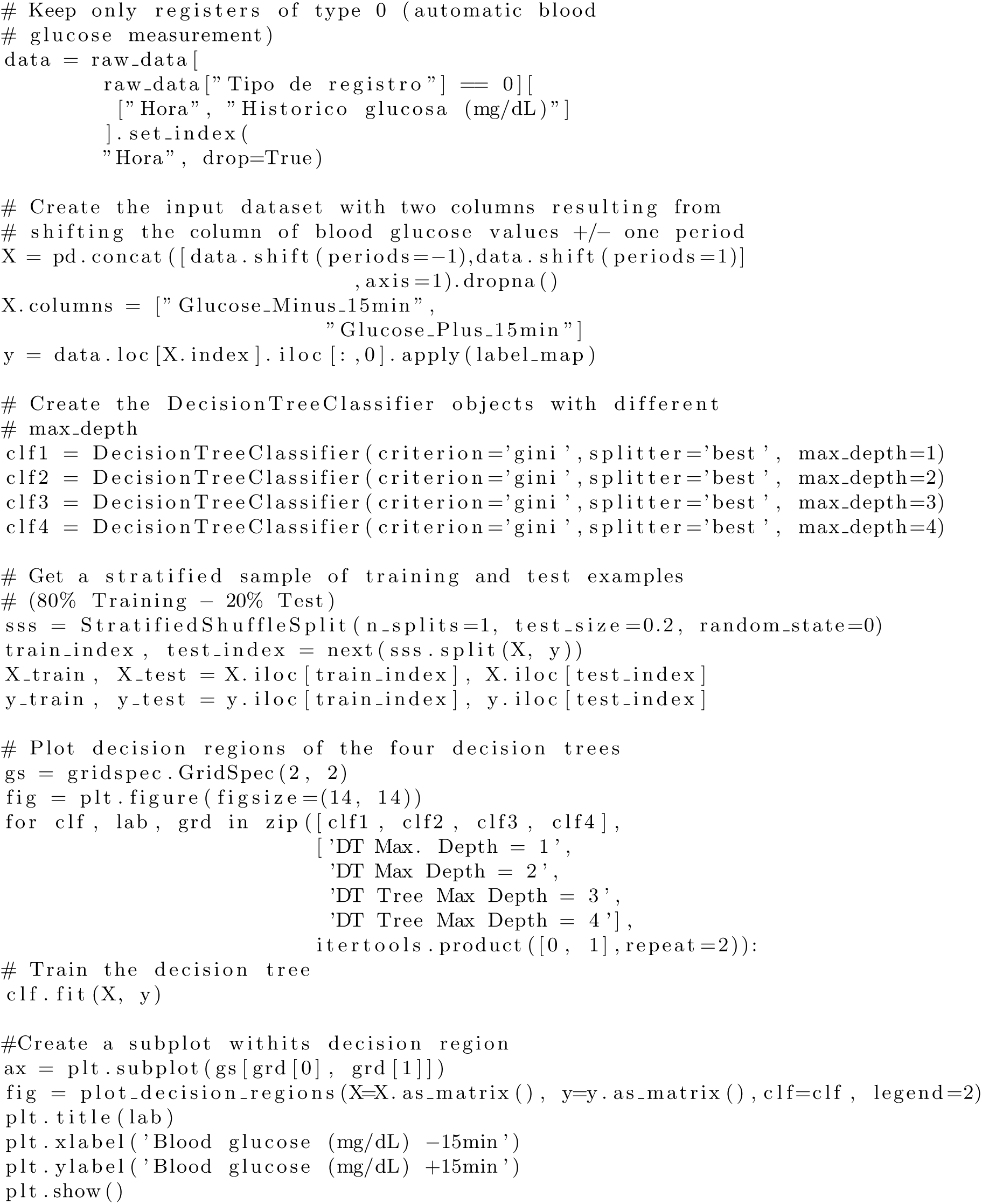

## B Report example

### Report of patterns

ACN

31/03/16 - 14/04/16

### Hyperglycemia patterns

#### Pattern 1

- Maximum level of glucose of the previous block is lower or equal than 234.5
- Mean level of glucose of the previous block is lower or equal than 145.3
- Maximum level of glucose of the previous day is greater than 192
- Standard deviation of the level of glucose of the previous block is lower or equal than 40.99
- Standard deviation of the level of glucose of the previous day is lower or equal than 40.95

Samples: 210 (10.43%)

Impurity: 0

Number of positive samples: 210 (25.93%)

Number of negative samples: 0 (0.00%)

#### Pattern 2

- Maximum level of glucose of the previous block is greater than 234.5

Samples: 210 (10.43%)

Impurity: 0.2024

Number of positive samples: 186 (22.96%)

Number of negative samples: 24 (2.00%)

### Hypoglycemia patterns

#### Pattern 1

- Maximum level of glucose of the previous block is greater than 139 and is lower or equal than 208.5
- Glycemic variability (MAGE) of the previous day is greater than 81.53
- Mean of the level of glucose of the previous day is lower or equal than 121.1

Samples: 297 (14.75%)

Impurity: 0.271

Number of positive samples: 249 (18.11%)

Number of negative samples: 48 (7.52%)

#### Pattern 2

- Maximum level of glucose of the previous block is greater than 139 and is lower or equal than 208.5
- Glycemic variability (MAGE) of the previous day is greater than 81.53
- Mean of the level of glucose of the previous day is greater than 121.1

Samples: 519 (25.78%)

Impurity: 0

Number of positive samples: 519 (37.75%)

Number of negative samples: 0 (0.00%)

### Decision trees

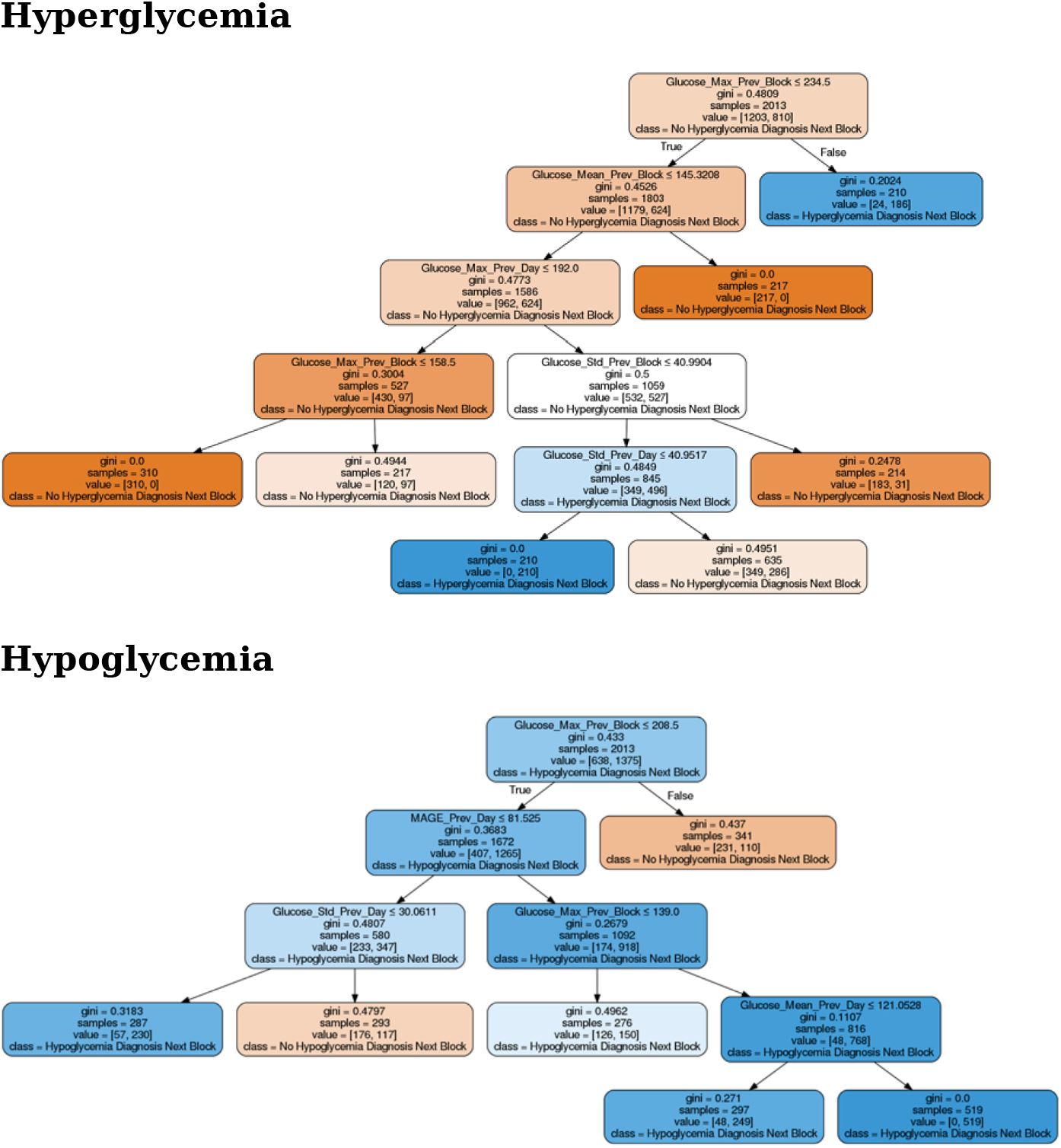
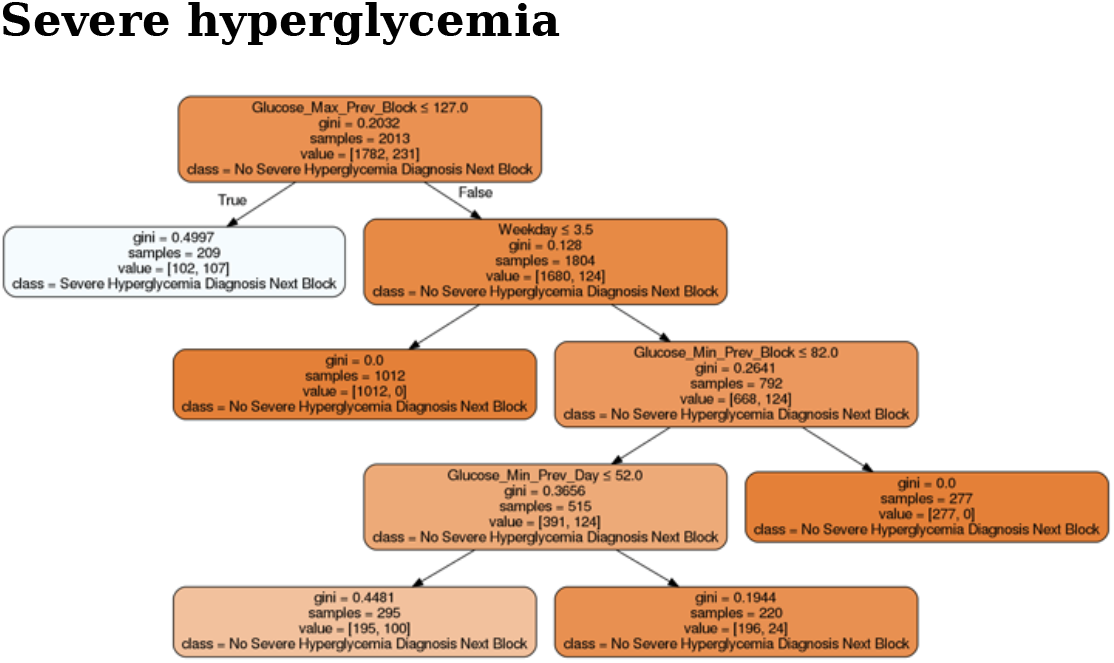

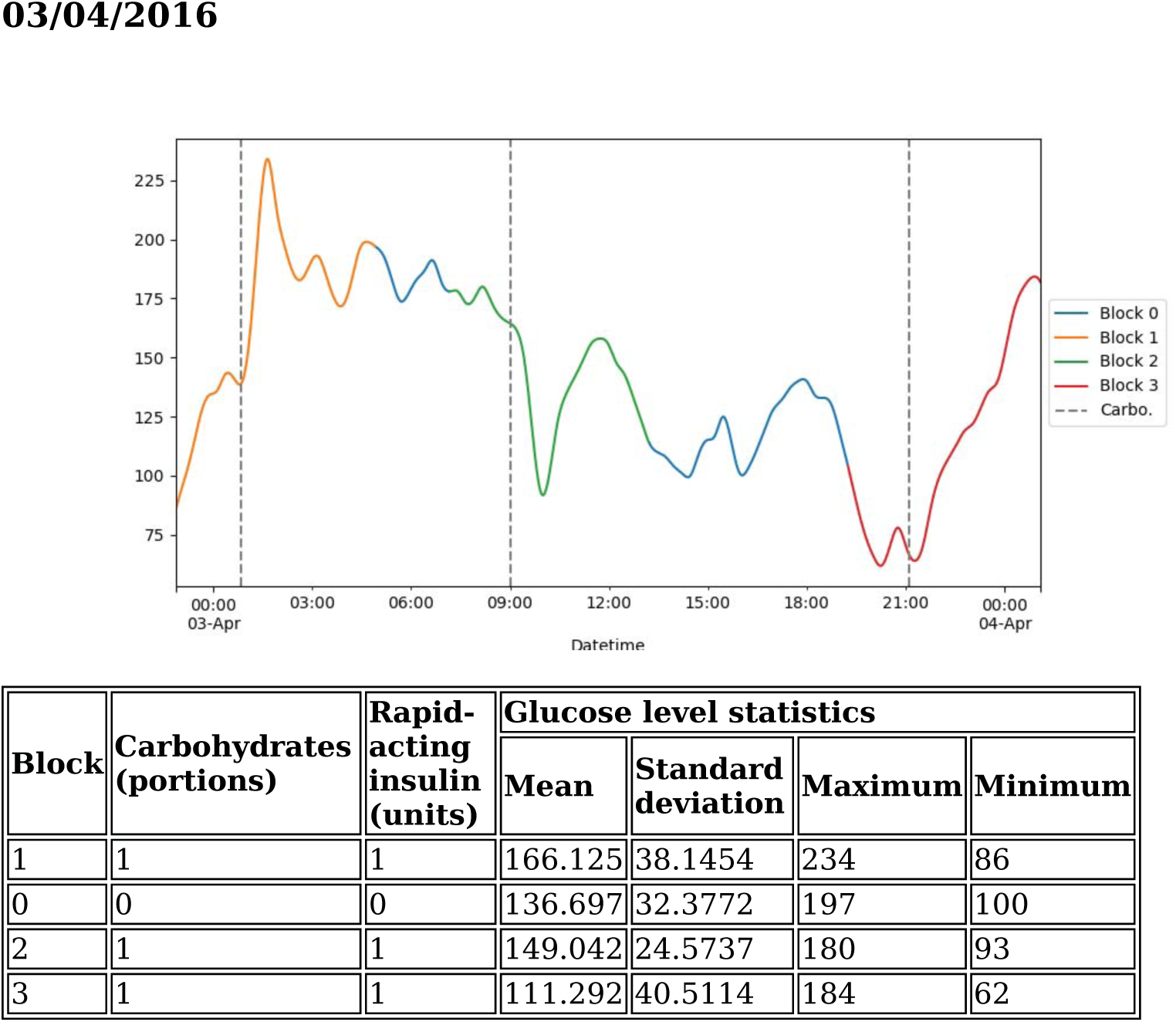

#### Day summary of glucose values

- Mean of the level of glucose of the day: 140.438
- Standard deviation of the level of glucose of the previous day: 38.8674
- Maximum level of glucose of the previous day: 234
- Minimum level of glucose of the previous day: 62
- Glycemic variability (MAGE): 76.4

## Report information

### What is a pattern?

A pattern is a set of rules that describe a situation in the patient’s life using several metrics related to his glucose level, insulin values, carbohydrates and time variables. Through these rules, it is possible to identify risk situation with regard to the glucose levels of the patient.

### What is a block?

Each day of the patient is divided in a series of blocks defined by the different meals that the patient has throughout the day. One block is defined by a time period that starts two hours before a meal and finishes four hours later. The blocks may be overlapped and include intakes of carbohydrates and insulin doses that are present also in other blocks. Block zero always corresponds to the remaining time that is not framed by any other block

### What type of patterns are there?

The information that a pattern provides depends on its type. The patterns can be of three types: Hyperglycemia, hypoglycemia and severe hyperglycemia patterns. The patterns of each type describe situations when the patient suffers from a disorder in his glucose level in the next meal block. The thresholds that determine a risk situation are the following ones:

- Hyperglycemia: 180 mg/dL
- Hypoglycemia: 70 mg/dL
- Severe hyperglycemia: 240 mg/dL

### What information does a pattern provide?

Each pattern is composed by the following elements:

1. Set of rules that describe the risk situation in the next block
2. Number of sumples of the data set that are obtained by the defined rules in the pattern. Moreover, it is displayed the percentage of samples regarding to the total of the data set
3. Impurity of the pattern using Gini impurity. It reaches its minimum (zero) when all the samples obtained by the defined rules are classified as risk situation in the next block
4. Number of positive samples, classified as a risk situation in the next block. Moreover, it shows the percentage of samples regarding to the total of positive samples
5. Number of negative samples, classified as a situation without risk in the next block. Moreover, it shows the percentage of samples regarding to the total of negative samples

### How to assess a pattern?

To assess a pattern it is necessary to consider two factors:

1. The number of samples must be relevant. The threshold of minimum samples is a 10% of the total, but this does not mean that it is the minimum number to consider a pattern as relevant. This criteria must be set by the patient or the physician.
2. The pattern does not have to have a minimal impurity. Long patterns with complex rules tend to overfit the model even though the maximum depth of the tree is set to 5 levels. This means that, although the pattern’s impurity was close to zero, the model would be overfitted and not be accurate in future risk situations.

### How to interpret a decision tree?

The decision tree contains both the patterns of a risk situation and the patterns of a situation with no risk. The nodes of the tree can be interpreted through their written information or their color. The color scale of each node determines both the majority class of the samples obtained by the split made in the parent node and how impure is the split. In the following color scale, it can be observed the impurity of the split according to the color intensity of each node:

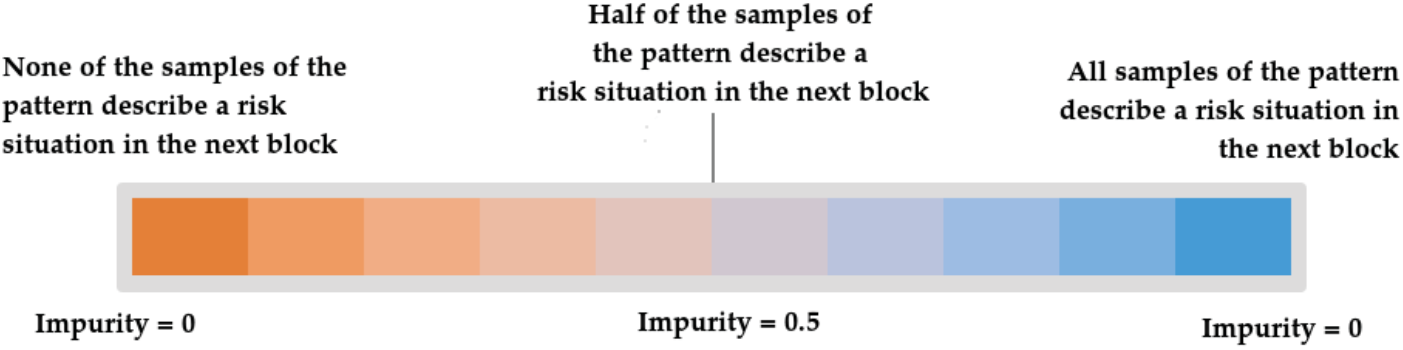

## C Examples of decision trees

The next decision trees were generated using the data file provided by the patient 3 during the period between 07/09/2016 and 22/09/2016. The rest of the reports of the patients, which contain the decision trees, are available at the following link of Google Drive: https://drive.google.com/drive/folders/0Bw6PbR_m3nxNUS1XZlIwQ21QWDg.

